# The fMRI signature of acute catatonic state and its response to benzodiazepines

**DOI:** 10.1101/2021.03.23.21253765

**Authors:** Pravesh Parekh, Anirban Gozi, Venkata Senthil Kumar Reddi, Jitender Saini, John P. John

**Affiliations:** Multimodal Brain Image Analysis Laboratory, National Institute of Mental Health and Neurosciences, Bangalore - 560029, India; Department of Psychiatry, National Institute of Mental Health and Neurosciences, Bangalore - 560029, India; Department of Neuroimaging and Interventional Radiology, National Institute of Mental Health and Neurosciences, Bangalore - 560029, India

**Keywords:** catatonia, lorazepam, neuroimaging, cortical complexity, resting state

## Abstract

We report for the first time, functional MRI markers of the acute retarded catatonic state and its response to benzodiazepines.

In this cross-sectional MRI study, we have compared the resting state whole-brain, within-network and seed (left precentral gyrus)-to-voxel connectivity, as well as cortical complexity between a sample of patients in acute retarded catatonic state (*n* = 15) and a demographically-matched healthy control sample (*n* = 15). Additionally, we examined whether the above variables were different between responders (*n* = 9) and non-responders (*n* = 6) to lorazepam.

Acute retarded catatonia was characterized by reduced functional connectivity, most robustly within the sensorimotor network, diffuse long-range hyperconnectivity, and seed (left precentral gyrus)-to-voxel hyperconnectivity in the frontoparietal and cerebellar regions. The lorazepam responders showed long-range as well as seed-to-voxel functional hyperconnectivity in comparison to the non-responders. Seed (left precentral gyrus)-to-voxel connectivity was positively correlated to the catatonia motor ratings. The catatonia sample showed a cluster of reduced vertex-wise cortical complexity in the right insular cortex and contiguous areas.

We have identified neuroimaging markers that characterize the acute retarded catatonic state and predict treatment response. We discuss how these novel findings have important translational implications for understanding the pathophysiology of catatonia and for predicting treatment response to benzodiazepines.

## Introduction

Catatonia is a complex syndrome of severe psychomotor dysregulation seen in various neuropsychiatric conditions.^1,2^ Irrespective of the nature of the underlying primary neuropsychiatric disorder, the acute retarded catatonic syndrome shows a remarkable similarity in clinical presentation across all patients suffering from the condition. Moreover, it is a unique psychiatric syndrome that dramatically responds fully to acute treatment in a matter of hours to days, and therefore offers a very promising disease model for studying neuroimaging markers that predict treatment response. However, functional MRI (fMRI) studies in catatonia have so far been reported only in patients following recovery, or in stable patients having mild catatonic symptoms^2,3^ (see Supplementary Table 1). This is likely due to the immense challenges involved in acquiring fMRI scans in the short time window between clinical presentation of the patient to the emergency psychiatry services and recovery from catatonia following initiation of treatment.

We aimed to study structural and resting state fMRI abnormalities in patients with acute retarded catatonia presenting to the psychiatric emergency services. We compared resting state functional connectivity (rs-FC) between the acute retarded catatonia sample and a demographically matched healthy sample; and attempted to link these with treatment response to benzodiazepines. There have been no published resting fMRI studies in *acute* catatonia so far^2^ (refer Supplementary Table 1 for a summary of previous functional brain imaging studies in catatonia). Nonetheless, since acute retarded catatonia is a condition typically characterized by severe psychomotor retardation but somewhat paradoxically responds dramatically to the GABA-ergic agonists, benzodiazepines, we hypothesized that the acute catatonia sample will have higher rs-FC in comparison to the healthy control sample. We also explored the differences in rs-FC between responders to benzodiazepines and non-responders, and the relationship between seed-to-voxel rs-FC from the left precentral gyrus (PCG) and severity of motor symptoms. Structural imaging studies in catatonia have reported inconsistent findings (for review, see).^2^ A measure of cortical folding complexity known as cortical surface complexity^4^ capable of detecting relatively small local differences, has been proposed as a temporally stable neurodevelopmental marker of cortical shape and abnormal gyrification in various neuropsychiatric disorders.^4–9^. We therefore examined alterations in regional brain volumes (using voxel-based morphometry, VBM) and surface complexity in our sample of patients with acute retarded catatonia in comparison to the healthy control sample.

## Materials and methods

### Study samples

In this first fMRI study in acute retarded catatonia, we recruited a sample of 15 right-handed patients (8 female patients) from the psychiatric emergency services of the National Institute of Mental Health and Neurosciences (NIMHANS), Bangalore, India, after obtaining written informed consent on behalf of the patients from the accompanying caregiver/legally authorized representative (following recovery from catatonia, written informed consent was obtained from patients as well) with the full approval of the NIMHANS Ethics Committee. Within this sample of patients, nine patients responded to lorazepam (‘LZM’ subgroup) while the remaining six did not; and required electroconvulsive therapy (ECT) for resolution of their catatonia state (‘ECT’ subgroup). We additionally recruited an age-matched sample of 15 right-handed healthy subjects (8 female subjects; ‘HS’ group). A detailed description of patient recruitment, establishment of diagnoses as per Diagnostic and Statistical Manual (DSM)-5^10^ criteria, socio-demographic details (Supplementary Table 2), and subject-wise clinical details (Supplementary Table 3) is provided in the Supplementary material.

### MRI Data acquisition

Structural brain imaging (CT/MRI) is an integral part of the clinical evaluation of patients who present to the psychiatric emergency services with catatonia.^11^ We aimed at acquiring the MRI scan while the patients were in the acute retarded catatonic state, and wherever possible, prior to initiation of treatment for catatonia. For seven out of 15 patients, we were able to schedule the MRI acquisition promptly before the patients were initiated on lorazepam; for the remaining eight patients, we were able to perform the MRI acquisition only after patients were initiated on treatment with lorazepam, typically due to scanner unavailability at a short notice. The initiation of treatment was never delayed on account of the patient’s participation in the study. The baseline Bush Francis Catatonia Rating Scale^12^ (BFCRS) rating was performed within an hour prior to the MRI acquisition for all the patients (mean score: 21.07 ± 5.69; range: 12-31; Supplementary Tables 2 and 3), indicating that all patients were in acute retarded catatonic state at the time of MRI acquisition.

The MRI of the patients and healthy subjects (except the last four healthy subjects—see below) were acquired on a 3T Philips Achieva scanner and consisted of a high-resolution T1-weighted scan and an eyes-open resting-state echo planar imaging (EPI) blood oxygenation level-dependent (BOLD) fMRI scan. The data of the last four (out of 15) healthy participants were acquired on a 3T Philips Ingenia CX scanner with a slightly different set of acquisition parameters. This was necessitated due to a scheduled hardware upgradation of the Achieva scanner, before data acquisition for the present study could be completed (see Supplementary Table 4 for key acquisition parameters).

### Preprocessing, quality check, and analyses

We converted the Digital Imaging and Communications in Medicine (DICOM) images to The Neuroimaging Informatics Technology Initiative (NIfTI) format using dcm2niix^13^ v20181125 (https://github.com/rordenlab/dcm2niix). We reviewed the T1-weighted structural images to ensure that there were no gross anatomical abnormalities and MR artefacts. For functional images, we performed motion correction and then identified the time points with excessive motion; these time points were censored during the denoising step by modelling them as regressors. The two groups did not differ significantly in the total number of detected outliers. All analyses were performed on MATLAB R2016a (MathWorks, Natick, USA; https://www.mathworks.com). Structural images were processed using the Computational Anatomy Toolbox^14^ (http://dbm.neuro.uni-jena.de/cat; version 1727) with Statistical Parametric Mapping (SPM; https://www.fil.ion.ucl.ac.uk/spm; version 7771); functional images were processed using the default pipeline implemented in Conn functional connectivity toolbox^15^ version 18b with SPM version 7487 in the background (see Supplementary materials for preprocessing details, quality check, and analyses).

### Whole brain functional connectivity

For examining the whole-brain functional connectivity differences, we used the *atlas* parcellation scheme in Conn, consisting of 132 regions of interests (ROIs) (see Supplementary material for details and Supplementary Table 5 for a list of abbreviations for each ROI). Connectivity matrices for each subject were calculated as the Fisher transformed correlation coefficient between the hemodynamic response function (HRF)-weighted regional time series of these 132 ROIs. Then, we tested: a) whether there were any pairs of connections that showed significantly different mean connectivity between CAT and the HS samples; b) whether there were any pairs of connections that showed significantly different mean connectivity between the LZM and ECT subgroups. For each of these tests, we performed a two-sample two-tailed *t*-test with α=0.05, corrected for the number of regions using an FDR method (*p*-FDR seed level correction, in Conn terminology).

### Within network functional connectivity

We examined between-group differences in functional connectivity within the sensorimotor, salience, frontoparietal, cerebellar, and subcortical networks which encompass the critical brain regions that have been linked to hypokinetic catatonia.^3^ The sensorimotor, salience, and frontoparietal networks were defined using the *networks* atlas in Conn, while the cerebellar and subcortical networks were defined using the *atlas* parcellation scheme in Conn (see Supplementary Table 6 for ROIs in each network). For each of these networks, we computed the connectivity matrices and tested the same set of hypotheses as in the previous section. Since we performed a test for five networks separately, we applied an additional Bonferroni correction for multiple comparisons by testing the *p*-FDR values against a threshold of 0.01.

### Seed (left precentral gyrus)-to-voxel connectivity and regression analyses

Motor disturbances are considered by most authors as the central feature of catatonia.^3^ In order to examine the specific abnormalities of rs-FC of the primary motor cortex with other brain regions, we compared seed-to-voxel connectivity between the study groups with the left PCG as the seed (see Supplementary Fig. 1 for ROI definition). We tested the same set of hypotheses as in the whole-brain functional connectivity section using the same statistical threshold. We also performed a linear regression analysis to examine the relationship between the BFCRS motor sub-score on day one (baseline) and the left PCG-based seed-to-voxel connectivity. For this analysis, the day one motor score, which reflects the severity of the motor dysfunction at the time of fMRI acquisition, was the predictor of the seed-to-voxel connectivity; the statistical threshold was the same as before.

**Fig. 1.**
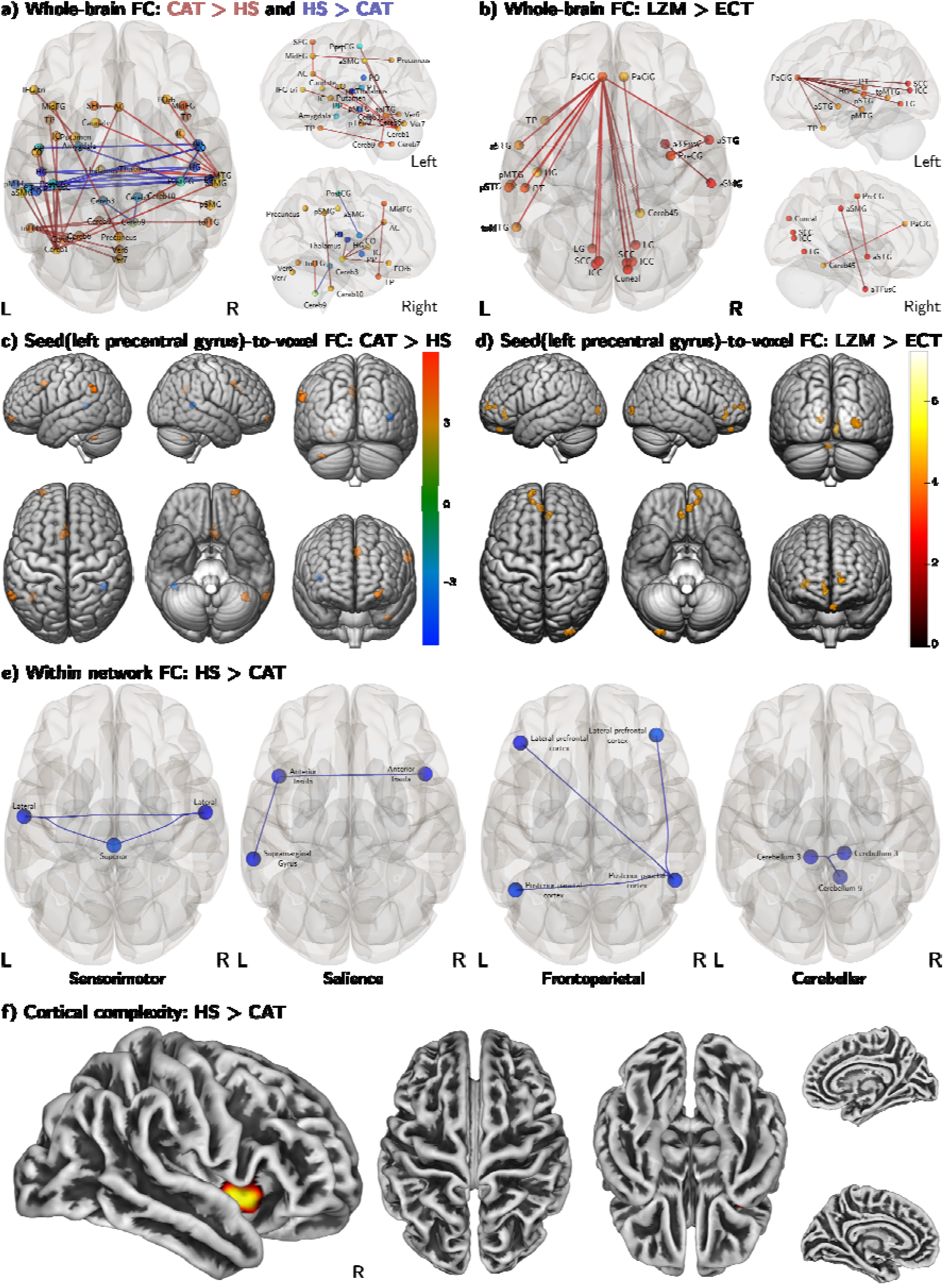
Altered resting state functional connectivity and cortical complexity in acute retarded catatonia (CAT) (*n* = 15) compared to healthy subjects (HS) (*n* = 15); and between the ‘lorazepam responders (LZM)’ (*n* = 9) and ‘lorazepam non-responders (ECT)’ (*n* = 6). (**A**) In this panel, we present the whole-brain FC differences between the CAT and HS samples at seed-level *p*-FDR < 0.05; we found widespread increased FC in long-range connections originating in the sensorimotor, salience, frontoparietal, temporal, and cerebellar regions; and decreased FC in the regional and/or interhemispheric connections of the sensorimotor, temporo-parietal and cerebellar regions. (**B**) In this panel, we present the whole-brain FC differences between the LZM and ECT samples at seed-level *p*-FDR < 0.05; we found increased FC in the LZM sub-group compared to the ECT sub-group. (**C**) In this panel, we present the differences in seed-to-voxel FC from the left precentral gyrus between the CAT and HS samples at voxel-wise *p*-uncorrected < 0.001 and cluster-wise *p*-FDR < 0.05^24^; we found four clusters of increased connectivity in the catatonia group; a single cluster of decreased connectivity noted in the figure was localized to the unlabelled region of the atlas. (**D**) In this panel, we present the differences in seed-to-voxel FC from left precentral gyrus between the LZM and ECT sub-samples at voxel-wise *p*-uncorrected < 0.001 and cluster-wise *p*-FDR < 0.05^24^; we found increased FC in the LZM sub-group as compared to ECT sub-group. (**E**) In this panel, we present the within-network FC differences of the sensorimotor, salience, frontoparietal, and cerebellar networks between the CAT and HS samples at seed-level *p*-FDR < 0.05 for each network; we found decreased FC within these networks in the catatonia group that comprised of brain regions which have previously been implicated in catatonia^3^; we did not observe any significant differences within the subcortical network between the two groups, nor were there any significant differences between the LZM and ECT sub-groups. (**F**) In this panel, we present the vertex-wise comparison of cortical complexity between HS and CAT samples (HS > CAT contrast) at *p* < 0.05, FWE corrected (using a non-parametric threshold-free cluster enhancement approach); we found lower cortical complexity in the catatonia group in a single cluster comprising predominantly the right insula and contiguous brain regions. There were no significant differences between the LZM and ECT sub-groups.

### Structural analyses

The resampled and smoothed cortical surface complexity files (see supplementary materials for details on processing) were entered into a general linear model framework. We tested the hypotheses whether the mean cortical surface complexity at each vertex was statistically different between the catatonia and the patient samples, and between the LZM and ECT subgroups. We used the threshold free cluster enhancement (TFCE) method^16^ as implemented in the TFCE toolbox (http://dbm.neuro.uni-jena.de/tfce; version 211) using default settings (5000 permutations) for estimating statistics and *p*-values. For the VBM analyses, we used the smoothed, modulated, normalized grey matter images with an absolute threshold masking of 0.1 and included the total intracranial volume as a covariate of no interest. We tested the same set of hypotheses as the cortical surface complexity analyses using the same TFCE approach.

## Reliability analyses

In order to estimate the generalizability of our results to the population, we performed reliability analyses of the between-group comparisons that tested our hypotheses (whole-brain and within-network rs-FC), using an iterative jackknife approach.^17^ We repeated these analyses by iteratively leaving out the data of one subject each from both catatonia and healthy groups. This gives us an estimate of the reliability of the results due to perturbations in the sample (see Supplementary material for details).

## Data availability

The data used in this study is available from the corresponding author, upon a reasonable request.

## Results

We observed widespread increased connectivity in the catatonia sample in the long-range connections from the sensorimotor, salience, frontoparietal, temporal, and cerebellar regions, while hypoconnectivity was noted in the regional and/or inter-hemispheric connections of the sensorimotor, temporo-parietal and cerebellar regions (Fig. 1, Supplementary Fig. 2, and Supplementary Table 7). The LZM subgroup showed significantly higher whole-brain connectivity between several pairs of regions, as compared to the ECT subgroup (Fig. 1, Supplementary Fig. 5 and Supplementary Table 9); most of these connections were from the left paracingulate region. The rs-FC results showed fair reliability (Dice coefficient: 0.60 ± 0.08), with eight pairs of connections showing consistent significant between-group differences across all jack-knife samples (see Supplementary material).

Within-network connectivity analysis revealed reduced connectivity in the catatonia sample in sensorimotor, salience, frontoparietal, and cerebellar networks (Fig. 1, Supplementary Fig. 6 and 7, and Supplementary Table 10), with no significant findings within the subcortical network or between the LZM and ECT subgroups. The Dice coefficients for reliability analysis of within-network functional connectivity differences between catatonia and healthy samples were ‘perfect’^17^ for the sensorimotor (1.00 ± 0.00), ‘high’^17^ for the frontoparietal (0.73 ± 0.12), and fair for the salience (0.63 ± 0.07) and cerebellar (0.62 ± 0.08) networks. All connections in the sensorimotor network, and multiple connections in the salience, frontoparietal and cerebellar networks were consistently different between the groups across all jack-knife samples. The sensorimotor network retained an excellent Dice coefficient (0.92 ± 0.15) indicating high reliability even after an additional Bonferroni correction for the five networks (Supplementary Fig. 8).

Left PCG seed-to-voxel functional connectivity analysis revealed four clusters of increased connectivity in the catatonia sample covering the frontoparietal and cerebellar regions (Fig. 1, Supplementary Fig. 10 and Supplementary Table 12). Similar to the whole brain connectivity results, significantly higher seed-to-voxel connectivity was seen in the LZM group covering the frontal and occipital regions (Fig. 1, Supplementary Fig. 12 and Supplementary Table 14). The left PCG seed-based connectivity showed a direct positive relationship with BFCRS motor sub-score (cluster size: 35 voxels; size *p*-FWE = 0.06, size *p*-FDR = 0.03; Supplementary Fig. 13).

We found a cluster of reduced vertex-wise cortical complexity in the catatonia sample in the right insular cortex and contiguous areas (*p* < 0.05 FWE corrected, TFCE, *p* = 0.01, 272 vertices in size) (Fig. 1, Supplementary Fig. 14). We did not find any volumetric differences in the VBM analysis (TFCE, *p* < 0.05, FWE corrected). Additionally, we did not find any differences between the LZM and ECT subgroups in VBM or cortical complexity analyses at this threshold.

## Discussion

Brain functional connectivity abnormalities have not been previously reported in acute catatonia. Here, we report reliable abnormalities of resting state functional connectivity in acute retarded catatonia that show a promising link with treatment response to benzodiazepines. The sensorimotor network comprising the primary motor cortex showed the most robust and reliable evidence for hypoconnectivity in acute retarded catatonia. Thus, the core pathophysiology in catatonia may involve a ‘chemical disconnection’ of the sensorimotor network and perhaps in the, salience, frontoparietal and cerebellar networks as well, that include critical brain regions linked to retarded catatonia.^3^ As an aberrant compensatory response to this regional hypoconnectivity, an abnormal long-range hyperconnectivity of these areas with other regions of the brain may ensue.^18^ This aberrant long-range functional hyperconnectivity, understandably mediated by an imbalance between the major excitatory and inhibitory neurotransmitters in the brain—glutamate and GABA— may cause the brain networks to go into a ‘psychomotor refractory state’ leading to the clinical features of acute retarded catatonia. A similar phenomenon has been reported, albeit at a different time scale, in epilepsy, where pathological high frequency oscillations (pHFO), which reflect inhibitory post-synaptic potentials from the GABA-ergic interneurons that regulate the glutamatergic pyramidal cell firing^19^ have been shown to be associated with a ‘cognitive refractory state’.^20^ The immediate therapeutic effect of benzodiazepines may only be on this aberrant hyperconnectivity (and not on the regional hypoconnectivity), as exemplified by how catatonic retardation re-emerges within 3-5 hours following a lorazepam challenge.^21–23^ The GABA-ergic agonistic action of benzodiazepines reverses the aberrant glutamate-mediated hyperconnectivity, and ‘releases’ the brain networks from the ‘psychomotor refractory state’, thereby resulting in a dramatic improvement of catatonic features. We did not observe any regional volumetric differences in our acute retarded catatonia sample without any neurological comorbidities. Reduced vertex-wise cortical complexity in the insular cortex, a critical brain region of the salience network, could be an interesting candidate for consideration as a temporally stable neurodevelopmental marker of cortical shape and abnormal gyrification^4^ in patients who are vulnerable for development of catatonia.

While the ‘psychomotor refractory state’ hypothesis of catatonia needs further validation, it provides a framework for understanding how benzodiazepines, somewhat paradoxically, produce marked clinical improvement in patients with acute retarded catatonia. In this initial study, we find preliminary evidence for how this aberrant hyperconnectivity is more pronounced in the lorazepam responder group in comparison to the lorazepam non-responder group. These novel findings have potential clinical applications in emergency settings to predict response to benzodiazepines and identify those patients who require early initiation of ECTs for recovery from acute retarded catatonia. Furthermore, this demonstration of the link between neuroimaging markers and treatment response in acute retarded catatonia may pave the way for similar functional neuroimaging studies, including pharmaco-imaging studies in other psychiatric disorders as well, which will potentially enhance our understanding of disease neurobiology and treatment.

## Data Availability

The data used in this study is available from the corresponding author, upon a reasonable request.

## Abbreviations

BFCRS: Bush Francis Catatonia Rating Scale
BOLD: blood oxygenation level-dependent
DICOM: Digital Imaging and Communications in Medicine
ECT: electroconvulsive therapy
EPI: echo planar imaging
fMRI: functional MRI
HRF: hemodynamic response function
HS: healthy subjects
LZM: lorazepam
NIfTI: The Neuroimaging Informatics Technology Initiative
NIMHANS: National Institute of Mental Health and Neurosciences
PCG: precentral gyrus
pHFO: pathological high frequency oscillations
ROI: region of interest
rs-FC: resting state functional connectivity
TFCE: threshold free cluster enhancement
VBM: voxel-based morphometry

## Acknowledgements

We acknowledge the support from NIMHANS for carrying out this research as part of the M.D. dissertation of A.G. We thank Mr. Krishnendu Vyas for assistance with the data acquisition of healthy subjects.

## Supplementary material

Supplementary material is available online.

## Author contributions

The author contributions are based on Contributor Roles Taxonomy (CRediT) (https://casrai.org/credit/):

**PP**: Methodology, Software, Validation, Formal analysis, Data Curation, Writing – Original Draft, Writing – Review & Editing, Visualization

**AG:** Conceptualization, Investigation, Data Curation, Writing – Review & Editing

**VSKR:** Conceptualization, Resources, Writing – Review & Editing, Supervision

**JS:** Conceptualization, Investigation, Resources, Writing – Review & Editing, Supervision, Project administration

**JPJ:** Conceptualization, Validation, Resources, Data Curation, Writing – Original Draft, Writing – Review & Editing, Visualization, Supervision, Project administration

