## Supplementary Materials for "The fMRI signature of acute catatonic state and its response to benzodiazepines"

#### --- *Supplementary Material* ---

Pravesh Parekh,[1,2,†] Anirban Gozi,[1,2,†] Venkata Senthil Kumar Reddi,[2] Jitender Saini,[3] and John P. John[1,2]

[†]**These authors contributed equally to this work.**

**Author affiliations:**

1 Multimodal Brain Image Analysis Laboratory, National Institute of Mental Health and Neurosciences, Bangalore - 560029, India

2 Department of Psychiatry, National Institute of Mental Health and Neurosciences, Bangalore - 560029, India

3 Department of Neuroimaging and Interventional Radiology, National Institute of Mental Health and Neurosciences, Bangalore - 560029, India

**Correspondence to:** John P. John

**Full address**: Multimodal Brain Image Analysis Laboratory,
National Institute of Mental Health and Neurosciences,
Bangalore – 560029,
India

### Summary of functional brain imaging studies in catatonia

**Table 1:** Summary of functional brain imaging studies in catatonia

| Study | Samples (*n*) | Catatonia status at the time of study | Neuroimaging modality/ies | Findings | Remarks |
|---|---|---|---|---|---|
| **Satoh et al., 1993[1]** | Catatonic subtype of SZ (*n* = 6) vs. other SZ (*n* = 13) and HS (*n* = 7) | 2-6 months after remission of catatonic symptoms | $^{123}$I IMP-SPECT | Reduced rCBF most prominent in bilateral parietal lobes and also in frontal regions | The patients with catatonia had been in remission for 2-6 months, and therefore, were not in an acute catatonic state at the time of the study; no catatonia ratings reported |
| **Northoff et al., 1999a[2]** | Akinetic catatonia (*n* = 10); psychiatric controls (*n* = 10; paranoid SZ *n* = 3; BPAD *n* = 7); HS (*n* = 20) | 8 days after full resolution of the akinetic catatonic syndrome following administration of lorazepam | $^{123}$I Iomazenil-SPECT and Tc-99mECD SPECT | Significantly lower Iomazenil binding in catatonia indicating decreased GABA-A receptor density in the left sensorimotor cortex; significantly lower rCBF in the right lower prefrontal and parietal cortices in catatonia | The imaging acquisition on the catatonia sample was performed following recovery from catatonia and the findings may indicate trait, and not state abnormalities |
| **Northoff et al., 1999b[3]** | 2 patients with akinetic catatonia | 1-2 hours after IV injection of 2 mg of Lorazepam | Task-based fMRI during repetitive sequential finger opposition (SFO) | Decreased motor activation in the left sensorimotor cortex during SFO task using the contralateral hand; reversal in laterality in the spatial extent of activated voxels during left-hand movements | This study on 2 patients with akinetic catatonia, 1-2 hours after Inj. Lorazepam did not show hemodynamic alterations in the SMA, indicating that the functional abnormalities may be confined to the primary motor cortex |
| **Northoff et al., 2000[4]** | Akinetic catatonia (*n* = 10); psychiatric controls (*n* = 10; paranoid SZ *n* = 3; BPAD *n* = 7); HS (*n* = 20) | 8 days after full resolution of the akinetic catatonic syndrome following administration of Lorazepam | Tc-99mECD SPECT and neuropsychological measures | Significantly lower rCBF in the right lower prefrontal and parietal cortices in catatonia; absence of correlation between parietal visuo-spatial abilities and right parietal rCBF in catatonia in contrast to psychiatric and healthy controls | The neuroimaging part of the study is the same as that reported in Northoff et al., 1999a[2] (see above) |

| | | | | | |
|---|---|---|---|---|---|
| **Escobar et al., 2000**[5] | Catatonia as per DSM-IV criteria ($N$ = 9; depression $n$ = 4; SZ $n$ = 5); no healthy comparison sample | Patients met criteria for catatonia during at least 2 consecutive weeks prior to enrolment; mean catatonia severity score: pre-ECT 20.8 (6.1); post-ECT 6.22 (6.43) on modified Rogers Scale (36-item scale) | rCBF using SPECT 1 week before first ECT and 1 week after last ECT (5-15 ECTs) | Significant increase in rCBF in parietal, temporal and occipital regions in patients with mood disorder following ECTs and not in patients with SZ | The sample comprises of patients who met criteria for catatonia during the course of at least 2 weeks prior to enrolment; during this period, the patients did not receive benzodiazepines |
| **Tiége et al., 2003**[6] | Case report of a 14-year-old girl with BPAD in an episode of severe depression and catatonia, with switch to mania following resolution of catatonia | PET-1 on day 2 in a drug-free state with acute catatonia (NCRS total score =19); PET-2 on day 8 following oral lorazepam treatment with patient having switched to mania | FDG-PET on day 2 and day 8 | PET-1 showed relative decrease in metabolism in anterior cingulate, medial prefrontal cortex, precuneus and dorsolateral cortices including left lateral parietal cortex, with relative hypermetabolism of the primary motor cortex, rostral part of the striatum and the vermis. PET-2 showed relative decrease of metabolism in the precuneus, lateral parietal cortices and right superior frontal gyrus | In this case report, hypermetabolism of the motor networks was observed during the catatonic state which was no longer noticed after resolution of catatonia and following switch to mania. Hypometabolism in the medial and lateral fronto-parietal areas persisted even after resolution of catatonia. It is difficult to make inferences based on a single case report, especially in the absence of corroboratory findings from group studies |
| **Northoff et al., 2004**[7] | Akinetic catatonia ($n$ = 10); psychiatric controls ($n$ = 10; paranoid SZ $n$ = 3; BPAD $n$ = 7); HS ($n$ = 10) | 8 days after full resolution of the akinetic catatonic syndrome following administration of lorazepam | Task-based fMRI during affective stimulation | Akinetic catatonic patients characterized by orbitofrontal cortical spatiotemporal alterations in negative and positive emotional processing | The final sample that entered into the analysis were 8 patients with catatonia and 7 psychiatric controls. The HS group had a mean age of 25.9 (6.1), as against the mean age of 41.6 (5.3) and 40.8 (4.9) of the catatonia and psychiatric control samples respectively (see Northoff |

| | | | | | |
|---|---|---|---|---|---|
| | | | | | et al., 1999a[2] and Northoff et al., 2000[4] above) |
| **Scheuerecker et al., 2009[8]** | Catatonic SZ (*n* = 12) vs HS (*n* = 12) | 1 month to 5 years after last catatonic episode (mean: 24.3 ± 22.8 months) | Task-based fMRI (self-initiated movements, externally triggered movements, rest) | Reduced activity during self-initiated movements in patients, compared to HS in right superior frontal gyrus, bilateral middle frontal gyrus, inferior frontal gyrus and parietal cortex | The patients with catatonic SZ were scanned 1 month to 5 years after the last catatonic episode |
| **Iseki et al., 2009[9]** | Case report of a 32-year-old left-handed male who presented with catatonic stupor, with acute aseptic encephalitis involving right frontotemporal area, with associated generalized convulsions and epilepsia partialis continua | Catatonic stupor | [$^{11}$C]-flumazenil PET; FDG PET; Tc-99 HMPAO SPECT; EEG; sMRI | Flumazenil PET during catatonic stupor showed decreased benzodiazepine receptor binding in the right frontotemporal area where glucose metabolism was preserved as revealed by FDG-PET. Reversal of abnormal right-sided anteriorly predominant cerebral hyperperfusion after injection of diazepam as noted using SPECT | The patient had generalized convulsions initially and epilepsia partialis continua for 2 weeks starting on the 23rd day after illness onset |
| **Richter et al., 2010[10]** | Akinetic catatonia (*n* = 6); HS (*n* = 8) | 6 weeks following remission from catatonia with administration of lorazepam | Task-based fMRI during affective stimulation following administration or lorazepam or placebo according to a random order in a double-blind design | Higher signal decreases in the OFC during negative stimuli after administration of lorazepam when compared to placebo in contrast to lower decreases in HS | The patients with akinetic catatonia were scanned 6 weeks following remission from catatonia. The authors interpret that signal decreases in patients with catatonia were regulated by lorazepam compared to HS |
| **Walther et al., 2017a[11]** | SZ (*n* = 42) sample stratified into those with catatonic symptoms (scoring >2 items on the BFCRS | The patients with SZ were recruited from inpatient and outpatient departments of a | Whole brain rCBF using ASL, and GM density using VBM | Higher perfusion in bilateral SMA in catatonia; increased catatonia was associated with higher perfusion in SMA. The catatonia sample had | The catatonia sub-sample was derived from a sample of 42 patients with SZ, recruited from the inpatient and outpatient departments using MINI and CASH interviews lasting |

| | | | | | |
|---|---|---|---|---|---|
| | (*n* = 15) and those without catatonia (*n* = 27); HS (*n* = 41) | university hospital through a MINI and the CASH. The mean BFCRS score of the catatonia sub-sample was 8.2 (5.2)]. 6 patients (2 with catatonia and 4 without) received benzodiazepines within 24 hours prior to MRI scanning | | lower GM density in frontal and insular cortices | at least 1 hour in total. Furthermore, the mean BFCRS score of the catatonia sample was 8.2 (s.d.=5.2), indicating that this is not an acute catatonia sample. Therefore, the neuroimaging findings may not reflect the acute catatonia state, but indicate important trait abnormalities in patients with SZ having catatonic symptoms |
| **Walther et al., 2017b**[12] | SZ (*n* = 46) and HS (*n* = 44) | Mean BFCRS score: 1.8 (3.6) | Resting state fMRI: ROI-ROI resting state functional connectivity | Catatonia and dyskinesia factor were correlated with thalamocortical connectivity; primary motor factor was correlated with connectivity between rostral anterior cingulate and caudate; spontaneous motor activity was correlated with connectivity between motor cortex and cerebellum in patients with SZ | This study was carried out in a sample of patients with SZ. This paper demonstrates the correlation between the motor symptom dimensions in SZ with motor networks; however, the SZ sample had a low mean BFCRS score and therefore, these are unlikely to reflect the state abnormalities of acute catatonia |
| **Foucher et al., 2018**[13] | SZ and schizoaffective disorders (*n* = 31) as per DSM-5 divided into 'cataphasia' (*n* = 9) and 'periodic catatonia' (*n* = 20) as per Wernicke-Kleist-Leonhard classification; HS (*n* = 27) | BFCRS score: mean (s.d.): periodic catatonia: 4.7 (3.0); cataphasia: 2.0 (2.6); benzodiazepine dose (diazepam equivalent-mg): periodic catatonia: 4.1 (6.7); cataphasia: 7.8 (20) | rCBF using ASL—'pure ASL' and 'ASL-BOLD' sequences | Increased rCBF in the left putamen and somatosensory cortex in the overall SZ sample. Periodic catatonia sample (*n* = 20) had higher rCBF than the HS and cataphasia samples in left precentral gyrus, posterior Broca's area, supplementary area and medial cingulate cortex. Cataphasia sample (*n* = 9) showed reduced rCBF than the HS and periodic catatonia | This study was carried out on stable patients with SZ and schizoaffective disorders, re-diagnosed into periodic catatonia and cataphasia, with a low catatonia severity score indicating that they were not in acute catatonia during the study |

| | | | | | |
|---|---|---|---|---|---|
| | | | | samples in the bilateral upper temporal gyrus and angular gyrus | |
| **Hirjak et al., 2020[14]** | Schizophrenia spectrum disorders (SSD) (*n* = 86) stratified into patients with catatonia (*n* = 24) and patients without catatonia (*n* = 22) on the basis of a cut-off score of 3 or more on the NCRS and at least 1 point in the 3 different symptom categories, i.e., motor, behavioral and affective | These patients have not had a history of acute catatonia. None of the patients were on benzodiazepines at the time of MRI and were on stable antipsychotic medications. Those who were on benzodiazepines were discontinued at least 72 hours before the MRI (the number of such patients are not specified) | Resting state fMRI and sMRI: intrinsic neural activity and GM volume | Predominantly frontothalamic and corticostriatal abnormalities in SSD patients with catatonia when compared to SSD patients without catatonia; corticostriatal and frontoparietal networks associated with catatonia affective scores; cerebellar and prefrontal cortical motor regions associated with catatonia behavioral scores | The SSD sample in this study comprised of patients who have not had a history of acute catatonia. The SSD sub-sample 'with catatonia' had a mean NCRS score of 6.88 (2.38) (maximum score = 80), indicating that the findings of the study do not reflect the 'state' abnormalities of acute catatonia |

**ASL**: arterial spin labeling; **BFCRS**: Bush Francis catatonia rating scale; **BOLD**: blood oxygenation level-dependent; **BPAD**: bipolar affective disorder; **CASH**: comprehensive assessment of symptoms and history; **DSM**: diagnostics and statistical manual of mental disorders; **EEG**: electroencephalography; **ECT**: electroconvulsive therapy; **FDG**: $^{18}$F-fluorodeoxy-glucose; **fMRI**: functional magnetic resonance imaging; **GM**: gray matter; **HS**: healthy subjects/healthy control sample; **MINI**: mini international neuropsychiatric interview; **NCRS**: Northoff catatonia rating scale; **OFC**: orbitofrontal cortex; **PET**: positron emission tomography; **rCBF**: regional cerebral blood flow; **ROI**: region of interest; **SMA**: supplementary motor area; **sMRI**: structural magnetic resonance imaging; **SPECT**: single-photon emission computed tomography; **SZ**: schizophrenia; **VBM**: voxel-based morphometry

### Materials and methods

#### Study samples

The study was conducted at the National Institute of Mental Health and Neurosciences (NIMHANS), Bangalore, India after obtaining permission from the Institute Ethics Committee. Patients in acute retarded catatonic state were recruited from the psychiatric emergency services of NIMHANS after obtaining written informed consent on behalf of the patients from the accompanying caregiver/legally authorized representative. Following recovery from catatonia, written informed consent was obtained from patients as well. The study samples were 15 right-handed patients in acute retarded catatonic state (henceforth referred as the 'CAT' group) and 15 age-, and gender-matched right-handed healthy comparison subjects (henceforth referred as the 'HS' group). The patients with acute catatonia were between the ages 18-40 years and did not have medical or neurological comorbidities that would have a significant influence on brain structure or function. We excluded participants with hyperkinetic/excited catatonia, comorbid psychoactive substance dependence other than nicotine or caffeine, as well as those having contraindications for undergoing MRI. Detailed physical examination and laboratory investigations including complete blood count, metabolic profile, serum CPK and serum Vitamin B12 were carried out for all patients as per the standard clinical protocol of management of catatonia. The diagnosis of catatonia and 'other psychiatric disorders' was made as per Diagnostic and Statistical Manual (DSM)-5 criteria[15] based on concordance between a trained clinician (A.G.) and the duty senior resident at the Emergency Psychiatry and Acute Care Services (EPAC) of NIMHANS, under the overall supervision of V.S.K.R. and J.P.J. A structured diagnostic assessment for 'other psychiatric disorders' was not incorporated in the study protocol at the time of recruitment, in view of the nature of the condition being studied, which renders the patients suffering from catatonia incapable of participating in a diagnostic interview prior to recruitment. Since the recruitment of participants was primarily from the psychiatric emergency services, and since at least some of the patients who responded promptly to lorazepam were expected to be discharged from the EPAC within two days without the need for further IP care, confirmation of the diagnosis of 'other psychiatric disorders' was achieved by reviewing the subsequent outpatient clinical evaluation notes of the respective treating units for those who responded promptly and were advised outpatient-based treatment ($n$ = 4); as well as clinical notes of the treating inpatient unit (a multidisciplinary team of post-graduate trainees and faculty from the departments of Psychiatry, Clinical

Psychology and Psychiatric Social Work) for those patients who underwent inpatient treatment (*n* = 11) (the medical records of all the patients who were recruited for this study are stored securely at the Medical Records Department of NIMHANS). A total of 18 patients with catatonia were recruited as part of the study, of whom the MRI data of one subject could not be retrieved due to technical glitches during data archival, while the data of two subjects were excluded due to poor quality structural images (see quality check section below). The consenting healthy comparison subjects did not have identifiable Axis-I psychiatric disorders; or medical/ neurological disorders that would have a significant influence on brain structure or function; or a history of major psychiatric disorders including substance dependence in first-degree relatives, as ascertained by a study-specific pro forma-based clinical interview.

All patients underwent standard treatment for catatonia and for the associated psychiatric and medical conditions under the respective clinical units at NIMHANS. The researchers did not have any role in treatment decisions but continued to administer the Bush Francis Catatonia Rating Scale[16] (BFCRS) daily to monitor the catatonia symptom severity (see below). Nine out of the 15 patients responded to lorazepam (henceforth referred as the 'LZM' subgroup) while the remaining six patients were non-responders and required electroconvulsive therapy (ECT) for the resolution of catatonia (henceforth referred as the 'ECT' subgroup). The severity of catatonia signs was quantified using the BFCRS[16] by a trained clinician (A.G.) after establishing good inter-rater reliability (intra-class correlation co-efficient for BFCRS severity score: 0.915) under the supervision of V.S.K.R. and J.P.J. The BFCRS severity score was computed by adding the score items 1-23.[16] We additionally computed a *motor sub-score* by adding the scores of the following items which were common between BFCRS and the Northoff Catatonia Rating Scale (NCRS),[17] from which the NCRS motor sub-score was computed: immobility/stupor, posturing/catalepsy, stereotypy, mannerisms, rigidity, waxy flexibility, gegenhalten, and ambitendency. The baseline BFCRS rating was performed within 1 hour prior to the MRI acquisition. Subsequently, daily ratings were carried out till the patient scored two or less on BFCRS, or till day 12 from baseline, whichever was earlier. The patients were deemed to have 'responded' to lorazepam or ECTs once they have achieved a BFCRS score of two or less, as they will no longer meet the diagnostic criteria for catatonia as per DSM-5 criteria.[18,19] Time to response was defined as the number of days required to reach a score of two or less following the baseline assessment (day one). A summary of the overall demographic and clinical variables of the CAT and HS samples is presented in **Table 2**. The clinical details of the catatonia sample including lorazepam responder status, age, gender,

DSM-5 diagnosis, medication status at baseline, duration of catatonia, overall duration of illness, BFCRS total score and motor sub-score, time to response and other relevant clinical details are given in **Table 3**.

**Table 2**: Summary of demographic and clinical variables of the acute retarded catatonia and healthy control samples

| Variable | Catatonia sample (*n* = 15) | | Healthy sample (*n* = 15) | Statistics* |
|---|---|---|---|---|
| Sex | 8 females, 7 males | | 8 females, 7 males | - |
| Age | 25.33 ± 6.03 (min = 18, max = 37) | | 27.27 ± 7.30 (min = 18, max = 41) | *T* (27.03) = -0.79; *p* = 0.44 |
| Education$ | 10.00 ± 3.05 (min = 4, max = 15) | | 16.00 ± 3.88 (min = 9, max = 24) | *T* (24.67) = -4.61; *p* < 0.00 |
| BFCRS score | Overall: 21.07 ± 5.69 | | - | - |
| | **LZM** 19.56 ± 5.75 (min = 12, max = 29) | **ECT** 23.33 ± 5.24 (min = 16, max = 31) | - | - |
| Motor sub-score# | Overall: 8.00 ± 2.73 | | - | - |
| | **LZM** 8.00 ± 2.73 (min = 3, max = 11) | **ECT** 8.00 ± 2.73 (min = 4, max = 12) | - | - |

* Two sample *t*-test statistics (two-tailed test assuming unequal variance; catatonia group > healthy group); $entry of education information for one participant in the healthy group was inadvertently missed in the socio-demographics sheet; reported values are excluding this missing information; #the mean and standard deviation of motor sub-score was the same between the lorazepam responders and non-responders

#### Magnetic Resonance Imaging (MRI) acquisition

We aimed at acquiring the MRI scan while the patients were in the acute retarded catatonic state, and wherever possible, prior to initiation of treatment for catatonia. For seven out of 15 patients, we were able to schedule the MRI acquisition promptly before the patients were initiated on lorazepam; for the remaining eight patients, we were able to perform the MRI acquisition only after patients were initiated on treatment with lorazepam (single dose of Inj. LZM 2 mg *n* = 5; two doses of Inj LZM 2 mg *n* =1; T. LZM 2 mg *n* = 2), typically due to scanner unavailability at a short notice.

During the resting state fMRI acquisition, the participants were given a standard instruction to relax without falling asleep and to not think of anything in particular, keeping their eyes open with gaze straight, and to avoid head and body movements (however, it was not typically possible to verify satisfactorily with the patient whether they understood this instruction owing to their catatonic state; typically, movement during scanning was not found to be substantial; the catatonia sample though, had a non-significantly higher average number of motion outliers than the healthy sample—see below). A summary of the pertinent image acquisition parameters is listed in **Table 4**.

**Table 3:** Subject-wise details of diagnosis, duration of illness, total BFCRS score, motor sub-score, and other clinical details; the first nine subjects are lorazepam responders (LZM subgroup) and the next six subjects are lorazepam non-responders (ECT subgroup); **BPAD**: bipolar affective disorder; **CPK:** creatinine phosphokinase; **ECT:** electroconvulsive therapy; **EPAC**: Emergency Psychiatry and Acute Care Services; **Inj.**: injection; **IV**: intravenous; **LZM**: lorazepam; **SGPT:** Serum glutamic pyruvic transaminase; **T.**: tablet; **THP**: trihexyphenidyl; **t.i.d.**: ter in die (thrice a day); **WNL**: within normal limits

| ID | Sex | Diagnosis (DSM-5) (Catatonia + other psychiatric disorders) | Medication status at recruitment | Duration of primary psychiatric illness (weeks) | Duration of catatonia (days) | Baseline BFCRS total | Baseline Motor sub-score | Time to response (days from baseline) | Remarks (if any) |
|---|---|---|---|---|---|---|---|---|---|
| ***Lorazepam responders*** | | | | | | | | | |
| sub-001 | Female | Unspecified schizophrenia spectrum and other psychotic disorder; with catatonia | Drug naïve | 8 | 3 | 20 | 10 | 2 | Tachycardia; Inj. LZM 2 mg IV stat given at EPAC ~1 hour prior to scanning, on account of delay in getting an MRI slot |
| sub-002 | Female | Major depressive disorder; single episode; severe; with psychotic features; with catatonia | Drug naïve | 18 | 7 | 12 | 3 | 2 | |
| sub-003 | Female | Unspecified schizophrenia spectrum and other psychotic disorder; with catatonia | Drug-free | 20 | 10 | 26 | 11 | 2 | Hemoglobin: 11.7 g%; other blood investigations: WNL |
| sub-004 | Male | Unspecified schizophrenia spectrum and other psychotic disorder; with catatonia | Drug naïve | 16 | 10 | 18 | 6 | 5 | Vitamin B12 and folate deficiency; 2 doses of Inj. Lorazepam 2 mg IV stat given at the EPAC ~6 hours and ~4 hours prior to scanning, on account of delay in getting an MRI slot |

| | | | | | | | | | |
|---|---|---|---|---|---|---|---|---|---|
| sub-005 | Female | Bipolar I disorder; current episode depression, severe; with psychotic features; with catatonia | On treatment with Quetiapine 600 mg and Lithium 900 mg till two days prior to MRI | 254 | 2 | 18 | 8 | 7 | Received 5 ECTs in the previous month, the last one 20 days prior to MRI |
| sub-006 | Male | Unspecified schizophrenia spectrum and other psychotic disorder; with catatonia | On treatment with Risperidone 6 mg and THP 2 mg | 9 | 6 | 12 | 5 | 2 | Mild increase in SGPT and S. Alkaline phosphatase; Inj. LZM 2 mg IV was given at EPAC~4 hours prior to scanning, on account of delay in getting an MRI slot |
| sub-007 | Female | Major depressive disorder; single episode; severe; with psychotic features; with catatonia | On treatment with Escitalopram 10 mg | 4 | 3 | 18 | 8 | 2 | Mild anemia; the patient was initiated on T. LZM 6 mg/day 3 weeks before, which was tapered and stopped, followed by onset of catatonia 3 days prior to recruitment; Inj. LZM 2 mg IV stat given at EPAC ~4 hours prior to scanning, on account of delay in getting an MRI slot |
| sub-008 | Female | Bipolar I disorder; current episode depression, severe; with psychotic features; with catatonia | Drug-free | 56 | 5 | 23 | 10 | 3 | Mild anemia |
| sub-009 | Male | Schizophrenia; continuous; with catatonia | On treatment with Risperidone 4 mg THP 4 mg | 60 | 180 | 29 | 11 | 2 | T. LZM 2 mg given ~8 hours prior to scanning, on account of delay in getting an MRI slot |

| | | | | | | | | | |
|---|---|---|---|---|---|---|---|---|---|
| | | | and T. LZM 4 mg/d | | | | | | |
| ***Lorazepam non-responders*** | | | | | | | | | |
| sub-010 | Male | Brief psychotic disorder; with catatonia | Drug naïve | 3 | 3 | 21 | 4 | 10 | Tachycardia, mildly elevated BP and CPK levels, Vit B12 deficiency and incontinence; investigations done to rule out autoimmune encephalitis |
| sub-011 | Male | Unspecified schizophrenia spectrum and other psychotic disorder; with catatonia | Drug-free | 520 | 4 | 31 | 12 | 8 | Tachycardia, high BP, mild fever and raised CPK which resolved after treatment with LZM; one dose of Inj. LZM 2 mg IV stat given at EPAC ~12 hours prior to scanning, on account of delay in getting an MRI slot; underwent ECT ~ 1 year prior to MRI; |
| sub-012 | Female | Unspecified schizophrenia spectrum and other psychotic disorder; with catatonia; Vit B12 deficiency | Drug naïve | 17 | 7 | 24 | 8 | 12 | Mild elevation of CPK; mild eosinophilia; Vitamin B 12 deficiency; Inj. LZM 2 mg IV stat given at EPAC ~8 hours prior to scanning, on account of delay in getting an MRI slot |
| sub-013 | Male | Unspecified schizophrenia spectrum and other psychotic disorder; with catatonia | Drug naïve | 17 | 20 | 21 | 8 | 7 | |
| sub-014 | Male | Schizophreniform disorder with good | On treatment with | 8 | 30 | 16 | 6 | 6 | Raised CPK; mildly raised plasma ammonia level; |

| | | | | | | | | | |
|---|---|---|---|---|---|---|---|---|---|
| | | prognostic factors; with catatonia | Risperidone 2 mg, THP 2 mg | | | | | | |
| sub-015 | Female | Schizophrenia; continuous; with catatonia; | Drug-free till three days prior to MRI | 208 | 365 | 27 | 10 | 15* | Mild anemia; Vitamin B12 deficiency; MRI could only be done three days after initiating T. LZM 2 mg HS, built up to 2 mg t.i.d. on the second day; T. LZM 2 mg given ~6 hours prior to MRI |

* This patient had not remitted as on day 12; the time to response was ascertained from the inpatient file notes

**Table 4:** Summary of key image acquisition parameters for T1-weighted scan (T1w) and resting state BOLD fMRI scan (rsfMRI) for Philips Achieva (*n* = 15 patients with catatonia and 11 healthy participants) and Philips Ingenia CX (*n* = 4 healthy participants)

| | T1w | | rsfMRI | |
|---|---|---|---|---|
| **Parameter** | **Achieva** | **Ingenia CX** | **Achieva** | **Ingenia CX** |
| Voxel size (mm) | 1.00 × 0.94 × 0.94 | 1.00 × 1.00 × 1.00 | 1.65 × 1.65 × 3.00 | 3.39 × 3.39 × 3.39 |
| Matrix size | 256 × 256 | 256 × 256 | 144 × 144 | 64 × 64 |
| Number of slices | 160 | 192[a] | 45 | 48 |
| TR (ms) | 8.11 to 8.36 | 6.51 to 6.52 | 2000 | 3000 |
| TE (ms) | 3.69 to 3.86 | 2.94 | 30 | 30 |
| Flip angle (degrees) | 8 | 9 | 90 | 90 |
| Number of volumes | - | - | 140 | 140 |

[a] One participant's data was acquired with 211 slices

#### Quality check

All the T1-weighted images were reviewed by an expert neuroradiologist and were opined to have no obvious structural abnormalities. Additionally, we visually examined the images for motion, fold-over, ghosting, susceptibility, and other MR-artefacts to ensure that these would not interfere with further processing and potentially impact the interpretation of the results. Data for two subjects in the catatonia group were discarded due to poor quality structural images on account of motion artifacts. For functional images, we performed motion correction (as part of the preprocessing steps; see below). We used the *97$^{th}$ percentile in normative sample* settings i.e., a global signal threshold of 5 and subject motion threshold of 0.9 mm, in Conn functional connectivity toolbox for identification of time points with excessive motion. These time points were censored during the denoising step by modelling them as regressors. In the catatonia group, the mean ± standard deviation of the number of detected motion outliers was 8.06 ± 12.70 (minimum = 0, maximum = 38), while, in the healthy group it was 2.25 ± 6.27 (minimum = 0, maximum = 25); although the number of motion outliers were higher in the catatonia group, the two groups did not differ significantly in the number of time points detected as outliers, as assessed by a two-tailed two-sample *t*-test assuming unequal variance [CAT>HS): $T$ (21.89) = 1.64, *p*-value = 0.12].

#### Structural preprocessing

The origin (0,0,0 coordinate) of T1-weighted structural images was approximately set to correspond to the anterior commissure using acpcdetect v2 (https://www.nitrc.org/forum/forum.php?forum_id=1927).[20–22] Images were then segmented into gray matter, white matter, and cerebrospinal fluid tissue classes using the Computational Anatomy Toolbox (CAT)[23] (http://dbm.neuro.uni-jena.de/cat; version 1727) with Statistical Parametric Mapping (SPM; https://www.fil.ion.ucl.ac.uk/spm; version 7771) in the background, running on MATLAB R2016a (MathWorks, Natick, Massachusetts, USA; https://www.mathworks.com). The modulated, normalized grey matter segmentation images were smoothed by a Gaussian kernel of 6mm at full width at half maximum. These images were used for voxel-based morphometry (VBM) analyses. The central surface files created during segmentation were then used for extracting cortical complexity.[24] Finally, the left and right hemisphere cortical complexity files were merged into a single mesh, resampled to a 32k mesh (Human Connectome Project) space, and smoothed by a filter of 20mm at full width at half maximum. All operations were performed using the tools available within CAT. For reporting the cortical complexity results, we applied a threshold of $p<0.05$ (FWE corrected) and used the Human Connectome Project multi-modal parcellation[25] for looking up the regions for clusters which were statistically significantly different between the groups (the full region names are based on the supplementary material provided in Glasser et al[25]).

#### Functional preprocessing and denoising of time series

Functional images were preprocessed using the default pipeline implemented in Conn functional connectivity toolbox[26] version 18b with SPM version 7487 in the background on MATLAB R2016a. The steps consisted of motion correction (using SPM's *realign and unwarp* method), centering i.e. setting the origin of the functional images to approximately correspond to the anterior commissure (translations only), detection of motion outliers (as mentioned before), segmentation and normalization of the functional images to the MNI space (normalization to a voxel size of 2 × 2 × 2 mm), and smoothing by a Gaussian kernel of 6 mm full-width at half maximum. In addition, centered structural images were also segmented and normalized; and white matter and cerebrospinal fluid masks were generated based on the segmentation. These masks were then twice eroded and added into the pipeline for denoising.

Conn implements an aCompCor[27] denoising approach; the time series of each subject was regressed to remove the effect of the following variables: the first five principal components derived from the white matter and cerebrospinal fluid masks (mentioned in the previous section), the six motion correction parameters (three translations, three rotations) and their first-order derivatives, the effect of motion outliers, and the main effect of resting state (which attempts to correct for 'ramping-up' effects at the beginning of the scan session). Additionally, we also performed a linear detrending of the time series. After regression, we used a band-pass filter of 0.008 – 0.09 Hz.

#### Specifying the regions of interest

For examining the whole-brain functional connectivity differences, we used the *atlas* parcellation scheme available in Conn; this scheme consists of 132 regions of interest (ROIs) where the cortical parcellation comes from the Harvard-Oxford maximum likelihood cortical atlas, the subcortical parcellation comes from the Harvard-Oxford maximum likelihood subcortical atlas, and the cerebellar parcellation comes from the automated anatomical labeling (AAL) atlas (please refer to the information provided in the Conn functional connectivity toolbox for a description of atlas construction and for original citations). A summary of the ROIs in this atlas along with the abbreviations used in the rest of this paper is presented in **Table 5**.

For within network connectivity differences, we defined the sensorimotor, salience, and frontoparietal networks using the *networks* atlas from Conn (this atlas is based on an independent component analysis performed in Conn on 497 subjects from the Human Connectome Project); we defined the cerebellar network using the cerebellar and vermis parcellations available from the *atlas* parcellation scheme in Conn; for the subcortical network, we selected the subcortical regions from the *atlas* parcellation scheme in Conn. The sensorimotor network consisted of three ROIs, the salience network consisted of seven ROIs, the frontoparietal network consisted of four ROIs, the cerebellar network consisted of 26 ROIs, and the subcortical network consisted of 10 ROIs (see **Table 6** for names of these regions).

For the seed (left precentral gyrus)-to voxel connectivity, we defined the left precentral gyrus from the *atlas* parcellation scheme from Conn (see **Figure 1** for visualization of this ROI).

**Table 5:** Abbreviations and full names of the regions of interest (ROIs) as defined in the ‘atlas’ parcellation scheme of the Conn functional connectivity toolbox; regions which have a left/right division are marked with an asterisk (*)

| Abbreviation | Full Name | Abbreviation | Full Name |
|---|---|---|---|
| ***Cortical Regions*** | | ***Cortical Regions (contd.)*** | |
| FP* | Fro. Pole | TOFusC* | Tem. Occipital Fusiform Cortex |
| IC* | Insular Cortex | OFusG* | Occipital Fusiform Gy. |
| SFG* | Sup. Fro. Gy. | FO* | Fro. Operculum Cortex |
| MidFG* | Mid. Fro. Gy. | CO* | Central Opercular Cortex |
| IFG tri* | Inf. Fro. Gy., pars triangularis | PO* | Parietal Operculum Cortex |
| IFG oper* | Inf. Fro. Gy., pars opercularis | PP* | Planum Polare |

| | | | |
|---|---|---|---|
| PreCG* | Precentral Gy. | HG* | Heschl's Gyrus |
| TP* | Tem. Pole | PT* | Planum Temporale |
| aSTG* | Sup. Tem. Gy., ant. division | SCC* | Supracalcarine Cortex |
| pSTG* | Sup. Tem. Gy., pos. division | OP* | Occipital Pole |
| aMTG* | Mid. Tem. Gy., ant. division | ***Subcortical Regions*** | |
| pMTG* | Mid. Tem. Gy., pos. division | Thalamus* | Thalamus |
| toMTG* | Mid. Tem. Gy., temporooccipital part | Caudate* | Caudate |
| aITG* | Inf. Tem. Gy., ant. division | Putamen* | Putamen |
| pITG* | Inf. Tem. Gy., pos. division | Pallidum* | Pallidum |
| toITG* | Inf. Tem. Gy., temporooccipital part | Hippocampus* | Hippocampus |
| PostCG* | Postcentral Gy. | Amygdala* | Amygdala |
| SPL* | Sup. Parietal Lobule | Accumbens* | Accumbens |
| aSMG* | Supramarginal Gy., ant. division | Brain Stem | Brain Stem |
| pSMG* | Supramarginal Gy., pos. division | ***Cerebellar Parcellations*** | |
| AG* | Angular Gy. | Cereb1* | Cerebellum Crus1 |
| sLOC* | Lat. Occipital Cortex, sup. division | Cereb2* | Cerebellum Crus2 |
| iLOC* | Lat. Occipital Cortex, inf. division | Cereb3* | Cerebellum 3 |
| ICC* | Intracalcarine Cortex | Cereb45* | Cerebellum 4 5 |
| MedFC | Fro. Med. Cortex | Cereb6* | Cerebellum 6 |
| SMA* | Juxtapositional Lobule Cortex# | Cereb7* | Cerebellum 7b |
| SubCalC | Subcallosal Cortex | Cereb8* | Cerebellum 8 |
| PaCiG* | Paracingulate Gy. | Cereb9* | Cerebellum 9 |
| AC | Cingulate Gy., ant. division | Cereb10* | Cerebellum 10 |
| PC | Cingulate Gy., pos. division | Ver12 | Vermis 1 2 |
| Precuneus | Precuneus Cortex | Ver3 | Vermis 3 |
| Cuneal* | Cuneal Cortex | Ver45 | Vermis 4 5 |
| FOrb* | Fro. Orbital Cortex | Ver6 | Vermis 6 |
| aPaHC* | Parahippocampal Gy., ant. division | Ver7 | Vermis 7 |
| pPaHC* | Parahippocampal Gy., pos. division | Ver8 | Vermis 8 |
| LG* | Lingual Gy. | Ver9 | Vermis 9 |
| aTFusC* | Temp. Fusiform Cortex, ant. division | Ver10 | Vermis 10 |
| pTFusC* | Temp. Fusiform Cortex, post. division | | |

**Ant.**: anterior; **Fro.**: frontal; **Gy.**: Gyrus; **Inf.**: inferior; **Lat.**: lateral; **Med.**: medial; **Mid.**: middle; **Pos.**: posterior; **Sup.**: superior; **Tem.**: temporal; #: formerly supplementary motor cortex

**Table 6:** List of regions subtended under sensorimotor, salience, frontoparietal, cerebellar, and subcortical networks; sensorimotor, salience, and frontoparietal networks are defined in the *networks* atlas of the Conn functional connectivity toolbox; cerebellar and subcortical networks were defined using the *atlas* parcellation scheme of the Conn functional connectivity toolbox; regions which have a left/right division are marked with an asterisk (*)

| Network | Regions |
|---|---|
| Sensorimotor | Lateral*; superior |
| Salience | Anterior cingulate; anterior insula*; rostral prefrontal cortex*; supramarginal gyrus* |
| Frontoparietal | Lateral prefrontal cortex*; posterior parietal cortex* |
| Cerebellar | Crus1*; crus2*; cerebellum 3*; cerebellum 4 5*; cerebellum 6*; cerebellum 7b*; cerebellum 8*; cerebellum 9*; cerebellum 10*; vermis 1 2; vermis 3; vermis 4 5; vermis 6; vermis 7; vermis 8; vermis 9; vermis 10 |
| Subcortical | Thalamus*; caudate*; putamen*; pallidum*; accumbens* |

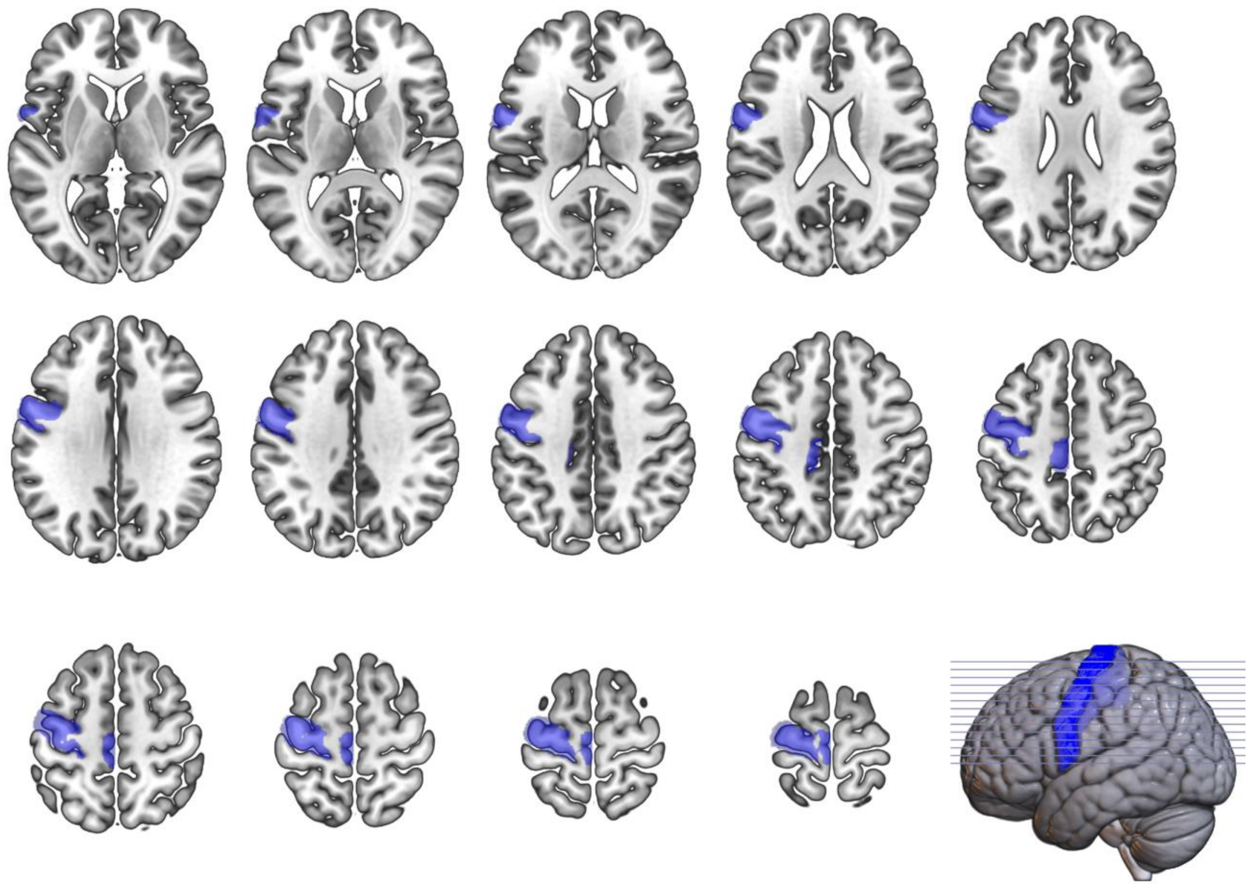

**Figure 1:** Extent of the left precentral gyrus as defined in the *atlas* parcellation scheme of the Conn functional connectivity toolbox; the image shows the span of the ROI (in blue color) overlaid on the spm152 template (from MRIcroGL); the axial slices range from +5 (top left) to +70 (bottom right) in increments of +5

#### Reliability analyses of the main results — Jackknife approach

In order to estimate the generalizability of our results to the population, we performed reliability analyses of the between-group comparisons that tested our hypotheses (whole-brain and within-network rs-FC), using an iterative jackknife approach.[28] We repeated these analyses by iteratively leaving out the data of one subject each from both catatonia and healthy groups. This gives us an estimate of the reliability of the results due to perturbations in the sample. Since we have images of 15 patients and 15 healthy subjects, there are 225 possible ways of creating subsets of 14 images in each group. For each of these subsets, we repeated the between group comparisons. Then, to summarize the results, we calculated the pairwise Dice similarity coefficient between the original results and the results obtained with the subset. In addition, we calculated the percentage of times (out of 225), that the same results as original was obtained. Such a strategy has previously been referred as "third-level fMRI analyses".[28] However, since our sample sizes were modest, we did not repeat the jackknife analyses by leaving out more than one subject each from the catatonia and healthy groups at a time. Additionally, given the large computational resources required for running non-parametric TFCE analyses, we did not perform jack-knife analyses for cortical complexity results.

#### Notes on visualization of results

For the visualization of cortical complexity results, we used the *cat_surf_results* function from the CAT toolbox (version 1753, available at https://www.jiscmail.ac.uk/cgi-bin/wa-jisc.exe?A2=ind2102&L=SPM&O=D&P=90329) for overlaying the log-transformed TFCE *p*-value images on a template. For the visualization of seed-to-voxel connectivity (including regression analysis) results, we present a multi-slice montage view generated by overlying the *T*-statistics map (after appropriate threshold; generated by Conn functional connectivity toolbox) on *spm152* template provided with MRIcroGL (v 1.2.20200331; https://github.com/rordenlab/MRIcroGL12). The lower limit of the color bar was set to zero (or if negative statistic value were present, then the minimum *T* value in this map) and the upper value was set to the maximum statistics value in this map. Depending on the direction of the effect, color scheme was set to one of: *winter* (for negative effects), *hot* or *warm* (for positive effects), and *blue2red* (for cases where both positive and negative effects were present). The selection of the axial slices was based on the peak of the significant clusters in each case. In certain cases, the *overlayDepth* setting in MRIcroGL was manipulated to make sure that the clusters were prominently visible in the rendered images (this only affects the rendered part of the image, not the multi-slice montage). The regions covered by these clusters was summarized as a figure based on the output lookup file generated by Conn functional connectivity toolbox; this 'lookup' is based on the atlas parcellation scheme (described previously). The color scheme for the lookup is *3-class Set 2* from ColorBrewer 2.0 (by Cynthia A. Brewer, Geography, Pennsylvania State University; http://colorbrewer2.org). The visualization of ROI-ROI connections (including the left, right, and superior view 3D displays) is from the Conn functional connectivity toolbox.

#### Note on non-labeled voxels

In some of the left precentral gyrus-based seed-to-voxel connectivity results, certain voxel clusters showing ‘significant’ between-group differences are marked as ‘not-labeled’; this is because the atlas used for labeling the voxels (*atlas* parcellation scheme which is part of the Conn functional connectivity toolbox) does not cover the entire brain. It is quite possible for some voxels at the edge of gray matter to show statistically ‘significant’ differences (partially owing to the smoothing step, as mentioned in the preprocessing section); however, such voxels may not be labeled in the atlas, as the cortical and subcortical parcellations in this atlas were created from Harvard-Oxford maximum likelihood atlas with a threshold of 25%. We have retained these voxels in the results for completeness.

#### Notes on calculation of effect size

We calculated the corrected Hedges’ $g$ as a measure of effect size, where the correction was applied to account for the small sample size[29]. For each computation, we calculated the pooled standard deviation as the square root of the weighted average of the squared group standard deviations, where the weighting was done by $n - 1$ (where $n$ is the group sample size).[29] Then, the corrected Hedges’ $g$ was calculated as[29]

$$g = \frac{\mu_1 - \mu_2}{SD_{pooled}} \times \left(\frac{N-3}{N-2.25}\right) \times \sqrt{\frac{N-2}{N}}$$

where $\mu_1$ is the mean of group one, and $\mu_2$ represents the mean of group two, $SD_{pooled}$ represents the pooled standard deviations, and $N$ represents the total sample size.

For ROI-ROI connectivity, we extracted the mean and standard deviations of the pairwise connections (which were significantly different between the groups). For seed-to-voxel connectivity, we calculated the mean and standard deviations using the *imcalc* utility of SPM for the clusters which showed a between group difference; we then extracted the mean and standard deviations of the peak voxel within the cluster for each group and then proceeded to calculate the effect size, as above. Irrespective of the direction of the effect, we have reported the absolute value of Hedges’ $g$.

### Results

#### Resting state functional connectivity abnormalities in acute catatonia

##### *Whole brain ROI-ROI connectivity: CAT (n = 15) > HS (n = 15)*

The whole brain ROI-ROI connectivity results are summarized in **Figure 2** and statistics between pairs of connections are listed in **Table 7**. The mean and standard deviation of the Dice coefficient from the jackknife reliability analysis was 0.60 ± 0.08; eight pairs of connections showed consistent significant between-group differences across all jackknife samples: the positive connections included connections between left cerebellum 7 and left temporal pole, left inferior frontal gyrus pars triangularis and left putamen, left putamen and left inferior frontal gyrus pars triangularis, and left temporal pole and left cerebellum 7; while negative connections included connections between left and right Heschl's gyrus, left Heschl's gyrus and right planum temporale, right planum temporale and left Heschl's gyrus, and right planum temporale and left parietal operculum cortex. See **Figure 3** for Dice coefficient for each jackknife sample.

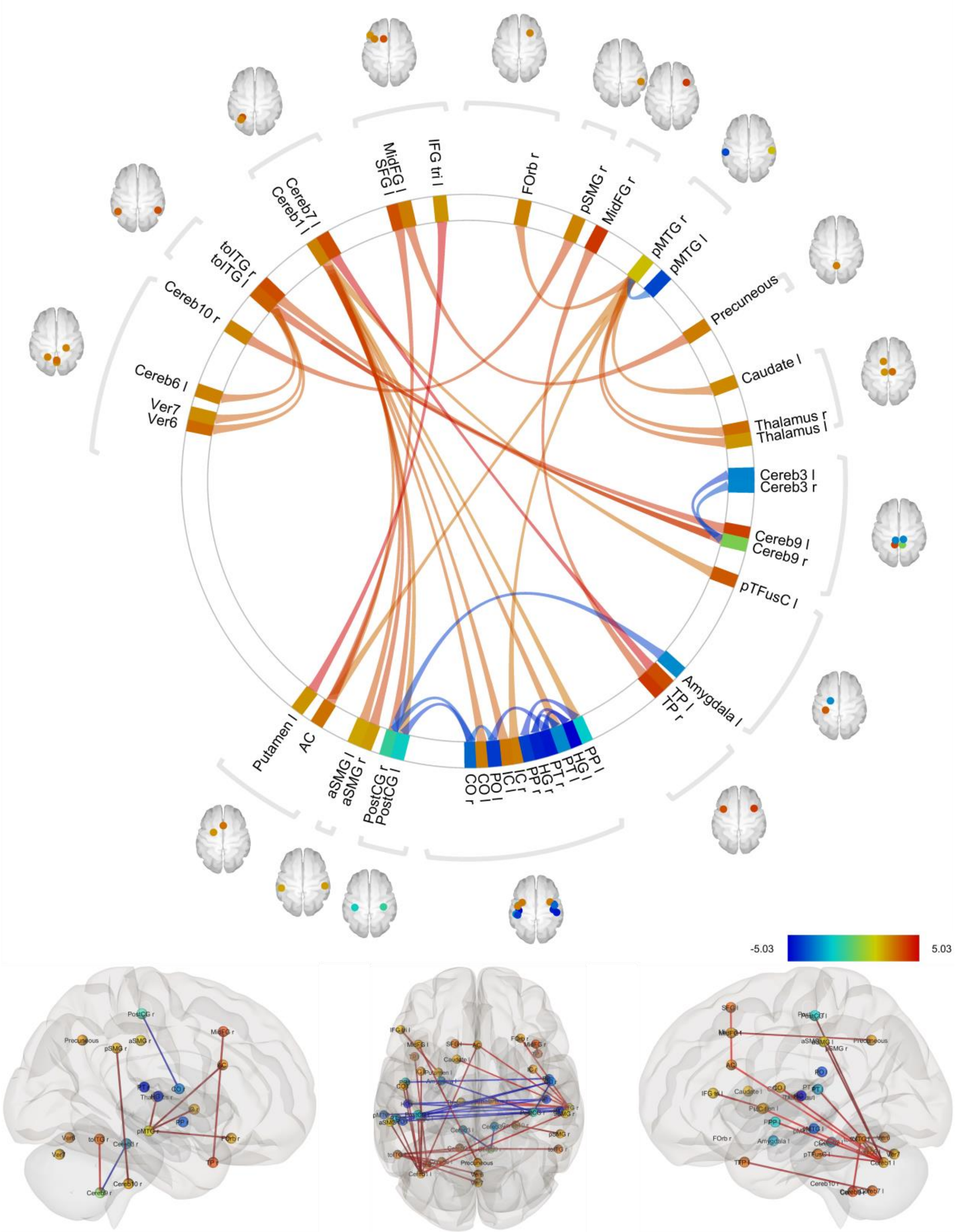

**Figure 2:** Comparison of pairwise ROI-ROI connectivity between catatonia ($n = 15$) and healthy ($n = 15$) groups (CAT > HS contrast) at seed-level $p$-FDR < 0.05 threshold; Fisher transformed correlation coefficient between the haemodynamic response function-weighted mean regional time series was used as a measure of connectivity; line colors correspond to the $T$-statistics (range: -5.03 to +5.03); see **Table 5** for expansion of the abbreviations used for the brain regions; an 'l' following the region name abbreviation indicates a region in the left hemisphere and 'r' following the region name abbreviation indicates a region in the right hemisphere

**Table 7:** Pairs of connections that were significantly different between catatonia group (*n* = 15) and healthy group (*n* = 15) (CAT > HS contrast) at seed-level *p*-FDR < 0.05 threshold; the first part of the table shows positive connections i.e. connections which were increased in the catatonia group as compared to the healthy group; and, the second part of the table shows negative connections i.e. connections which were reduced in the catatonia group as compared to healthy group; within each of these parts, the connections are ordered from anterior to posterior source ROIs, followed by subcortical source ROIs, and finally by cerebellar source ROIs; connections which were statistically significant in both directions have two values in the *p*-FDR column; *p*-values are rounded off to two decimal places; see **Table 5** for full form of the abbreviations of the names of the brain regions; an 'l' following the region name abbreviation indicates a region in the left hemisphere and 'r' following the region name abbreviation indicates a region in the right hemisphere

| Source | Target | *T* (df) Statistics | *p*-uncorrected | *p*-FDR | Hedges' *g* |
|---|---|---|---|---|---|
| **CAT > HS** | | | | | |
| IFG tri l | Putamen l | T (28) = 5.02 | < 0.00 | < 0.00 / < 0.00* | 1.72 |
| MidFG l | Precuneous | T (28) = 4.07 | < 0.00 | 0.05 / 0.05* | 1.40 |
| MidFG r | TP r | T (28) = 4.11 | < 0.00 | 0.04 / 0.04* | 1.41 |
| SFG l | AC | T (28) = 4.27 | < 0.00 | 0.03 / 0.03* | 1.47 |
| TP l | Cereb7 l | T (28) = 5.03 | < 0.00 | < 0.00 / < 0.00* | 1.73 |
| aSMG r | Cereb1 l | T (28) = 4.06 | < 0.00 | 0.05 / 0.03* | 1.39 |
| pMTG r | AC | T (28) = 3.36 | < 0.00 | 0.05 | 1.15 |
| pMTG r | Caudate l | T (28) = 3.49 | < 0.00 | 0.04 | 1.20 |
| pMTG r | FOrb r | T (28) = 3.78 | < 0.00 | 0.04 | 1.30 |
| pMTG r | IC r | T (28) = 3.30 | < 0.00 | 0.05 | 1.13 |
| pMTG r | Thalamus l | T (28) = 3.76 | < 0.00 | 0.04 | 1.29 |
| pMTG r | Thalamus r | T (28) = 3.74 | < 0.00 | 0.04 | 1.28 |
| PostCG r | Cereb1 l | T (28) = 4.00 | < 0.00 | 0.03 / 0.03* | 1.37 |
| pSMG r | Cereb10 r | T (28) = 4.06 | < 0.00 | 0.05 / 0.05* | 1.39 |
| toITG l | Cereb6 l | T (28) = 3.66 | < 0.00 | 0.03 | 1.26 |
| toITG l | Cereb9 l | T (28) = 4.32 | < 0.00 | 0.01 / 0.02* | 1.48 |
| toITG l | Cereb9 r | T (28) = 4.25 | < 0.00 | 0.01 / 0.01* | 1.46 |
| toITG l | Ver6 | T (28) = 3.57 | < 0.00 | 0.03 | 1.23 |
| toITG l | Ver7 | T (28) = 3.57 | < 0.00 | 0.03 | 1.23 |
| Cereb1 l | AC | T (28) = 3.62 | < 0.00 | 0.03 | 1.24 |
| Cereb1 l | aSMG l | T (28) = 3.74 | < 0.00 | 0.03 | 1.28 |
| Cereb1 l | CO l | T (28) = 3.63 | < 0.00 | 0.03 | 1.25 |
| Cereb1 l | IC l | T (28) = 3.57 | < 0.00 | 0.03 | 1.23 |
| Cereb1 l | PostCG l | T (28) = 3.32 | < 0.00 | 0.04 | 1.14 |
| Cereb1 l | PP l | T (28) = 3.24 | < 0.00 | 0.04 | 1.11 |
| Cereb1 l | PT l | T (28) = 3.53 | < 0.00 | 0.03 | 1.21 |
| Cereb1 l | pTFusC l | T (28) = 3.36 | < 0.00 | 0.04 | 1.15 |
| Cereb9 r | toITG r | T (28) = 3.91 | < 0.00 | 0.02 | 1.34 |
| **HS > CAT** | | | | | |

| | | | | | |
|---|---|---|---|---|---|
| PP l | PP r | T (28) = -4.52 | < 0.00 | 0.01 / 0.01* | 1.55 |
| CO r | PO l | T (28) = -3.69 | < 0.00 | 0.04 | 1.27 |
| CO r | PostCG l | T (28) = -4.19 | < 0.00 | 0.03 / 0.03* | 1.44 |
| CO r | PostCG r | T (28) = -3.81 | < 0.00 | 0.04 / 0.03* | 1.31 |
| HG l | HG r | T (28) = -4.60 | < 0.00 | 0.01 / 0.01* | 1.58 |
| HG l | PT r | T (28) = -4.71 | < 0.00 | 0.01 / 0.01* | 1.62 |
| HG r | PT l | T (28) = -4.09 | < 0.00 | 0.02 / 0.04* | 1.41 |
| PostCG r | Amygdala l | T (28) = -4.06 | < 0.00 | 0.03 / 0.05 | 1.39 |
| pMTG r | pMTG l | T (28) = -3.54 | < 0.00 | 0.04 | 1.21 |
| PO l | PT r | T (28) = -4.43 | < 0.00 | 0.02 / 0.01* | 1.52 |
| PT l | PT r | T (28) = -3.91 | < 0.00 | 0.04 / 0.02* | 1.34 |
| Cereb3 l | Cereb9 r | T (28) = -4.23 | < 0.00 | 0.03 / 0.01* | 1.45 |
| Cereb9 r | Cereb3 r | T (28) = -3.78 | < 0.00 | 0.02 | 1.30 |

*indicates connections which were statistically significant in both the directions (i.e., from Region A – Region B and Region B – Region A): the first *p*-value is for the reported connection and the second *p*-value is for the connection in the reverse direction

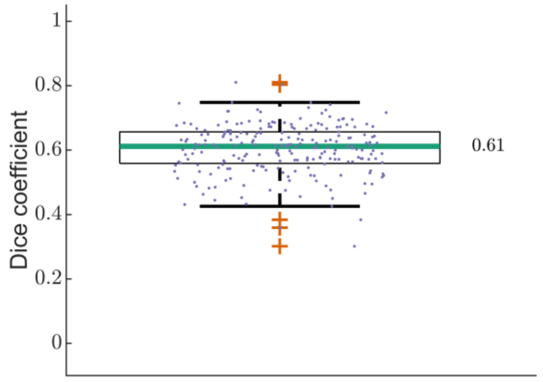

**Figure 3:** Boxplot of Dice coefficients from jackknife analyses for between group comparison of whole brain ROI-ROI connectivity (CAT > HS); the median Dice coefficient was 0.61

***Whole brain ROI-ROI connectivity: CAT (n = 15) > HS (Achieva only; n = 11)***

The overall number of pairs of brain regions showing significant difference in functional connectivity between healthy and catatonia samples was less in this analysis [CAT > HS (Achieva only) contrast] which excluded 4 healthy subjects whose images were acquired on the Ingenia CX scanner. However, the nature and direction of results were similar to the CAT > HS comparison done on the overall sample (see above). We found increased long-range connectivity and reduced cerebellar connectivity in the catatonia sample; these results are summarized in **Figure 4** and the statistics between pairs of connections are listed in **Table 8**.

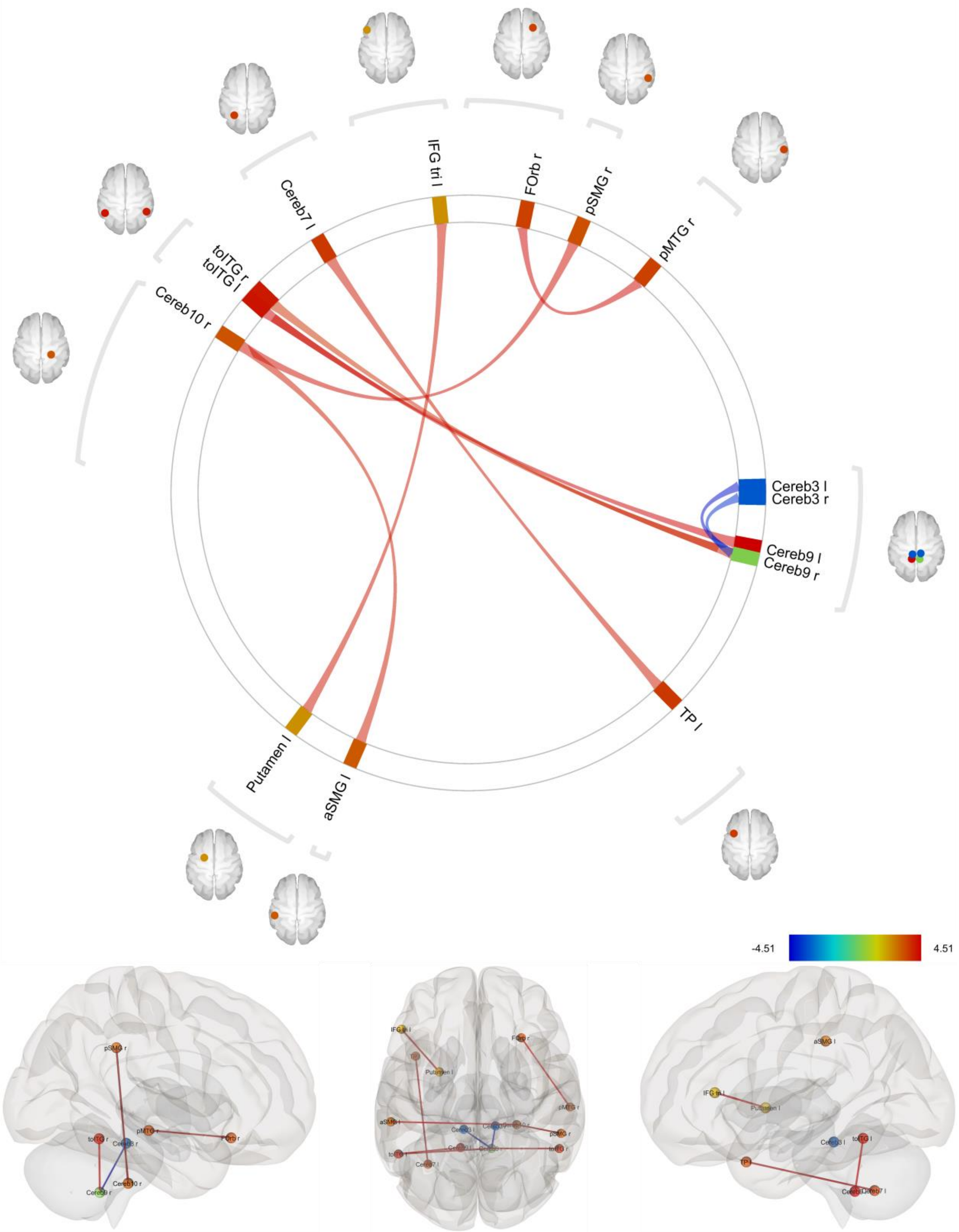

**Figure 4:** Results from comparison of pairwise ROI-ROI connectivity between catatonia group ($n$ = 15) and healthy groups (excluding those images which were acquired on Ingenia CX scanner; $n$ = 11) (CAT > HS (Achieva only) contrast) at seed-level $p$-FDR < 0.05 threshold; Fisher transformed correlation coefficient between the haemodynamic response function weighted mean regional time series was used as a measure of connectivity; line colors correspond to the $T$-statistics (range: -4.51 to +4.51); see **Table 5** for expansion of the abbreviations used for the brain regions; an ‘l’ following the region name abbreviation indicates a region in the left hemisphere and ‘r’ following the region name abbreviation indicates a region in the right hemisphere

**Table 8:** Pairs of connections that were significantly different between catatonia group (*n* = 15) and healthy group (excluding those images which were acquired on Ingenia CX scanner; *n* = 11) (CAT > HS (Achieva only) contrast) at seed-level *p*-FDR < 0.05 threshold; the first part of the table shows positive connections i.e. connections which were increased in the catatonia group as compared to the healthy group, and the second part of the table shows negative connections i.e. connections which were reduced in the catatonia group as compared to healthy group; within each of these parts, the connections are ordered from anterior to posterior source ROIs, followed by subcortical source ROIs, and finally by cerebellar source ROIs; connections which were statistically significant in both directions have two values in the *p*-FDR column; *p*-values are rounded off to two decimal places; see **Table 5** for full form of the abbreviations of the names of the brain regions; an 'l' following the region name abbreviation indicates a region in the left hemisphere and 'r' following the region name abbreviation indicates a region in the right hemisphere

| Source | Target | *T* (df) Statistics | *p*-uncorrected | *p*-FDR |
|---|---|---|---|---|
| **CAT > HS (Achieva only)** | | | | |
| IFG tri l | Putamen l | T (24) = 4.18 | < 0.00 | 0.04 / 0.04* |
| FOrb r | pMTG r | T (24) = 4.44 | < 0.00 | 0.02 / 0.02* |
| TP l | Cereb7 l | T (24) = 4.24 | < 0.00 | 0.04 / 0.04* |
| pSMG r | Cereb10 r | T (24) = 4.33 | < 0.00 | 0.03 / 0.03* |
| toITG l | Cereb9 l | T (24) = 4.51 | < 0.00 | 0.02 / 0.02* |
| toITG l | Cereb9 r | T (24) = 4.07 | < 0.00 | 0.03 / 0.03* |
| Cereb10 r | aSMG l | T (24) = 4.11 | < 0.00 | 0.03 |
| Cereb9 r | toITG r | T (24) = 3.81 | < 0.00 | 0.03 |
| **HS (Achieva only) > CAT** | | | | |
| Cereb3 l | Cereb9 r | T (24) = -4.32 | < 0.00 | 0.03 / 0.03* |
| Cereb9 r | Cereb3 r | T (24) = -3.78 | < 0.00 | 0.03 |

*indicates connections which were statistically significant in both the directions (i.e. from Region A – Region B and Region B – Region A): the first *p*-value is for the reported connection and the second *p*-value is for the connection in the reverse direction

##### *Whole brain ROI-ROI connectivity: LZM (n = 9) > ECT (n = 6)*

The whole brain ROI-ROI connectivity differences between lorazepam responders (*n* = 9) and lorazepam non-responders (*n* = 6) (LZM > ECT contrast), are summarized in **Figure 5** and detailed statistics are reported in **Table 9**.

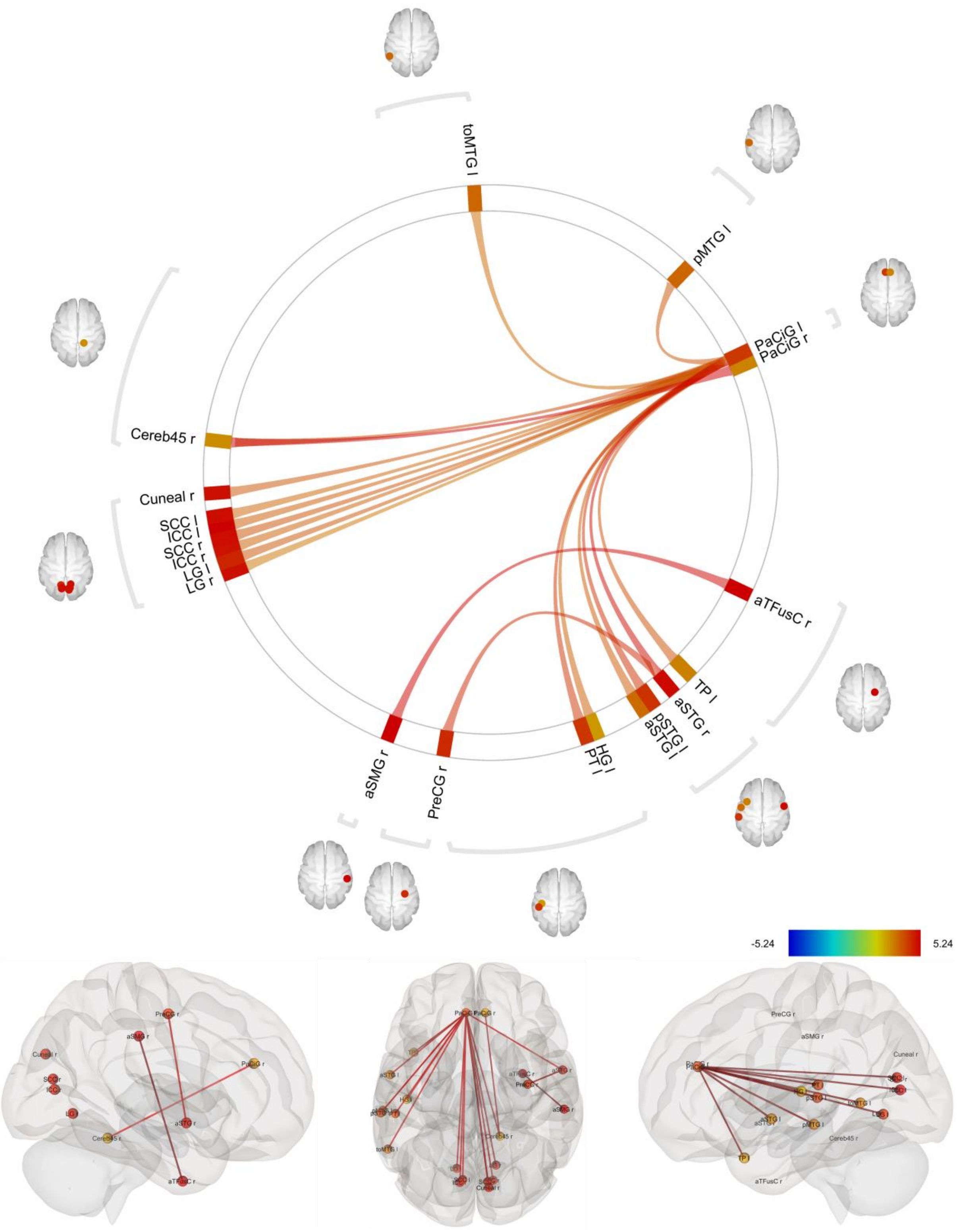

**Figure 5:** Comparison of pairwise ROI-ROI connectivity between lorazepam responder group ($n$ = 9) and lorazepam non-responder group ($n$ = 6) (LZM > ECT contrast) at seed-level $p$-FDR < 0.05 threshold; Fisher transformed correlation coefficient between the haemodynamic response function-weighted mean regional time series was used as a measure of connectivity; line colors correspond to the $T$-statistics (range: -5.24 to +5.24); see **Table 5** for expansion of the abbreviations used for the brain regions; an ‘l’ following the region name abbreviation indicates a region in the left hemisphere and ‘r’ following the region name abbreviation indicates a region in the right hemisphere

**Table 9:** Pairs of connections that were significantly different between lorazepam responder group ($n$ = 9) and lorazepam non-responder group ($n$ = 6) (LZM > ECT contrast) at seed-level $p$-FDR < 0.05 threshold; all connections were in the positive direction i.e. increased in the lorazepam responder group as compared to the lorazepam non-responder group; the connections are ordered from anterior to posterior source ROIs, followed by subcortical source ROIs, and finally by cerebellar source ROIs; connections which were statistically significant in both directions have two values in the $p$-FDR column; $p$-values are rounded off to two decimal places; see **Table 5** for full form of the abbreviations of the names of the brain regions; an 'l' following the region name abbreviation indicates a region in the left hemisphere and 'r' following the region name abbreviation indicates a region in the right hemisphere

| Source | Target | *T* (df) Statistics | *p*-uncorrected | *p*-FDR | Hedges' *g* |
|---|---|---|---|---|---|
| PaCiG l | TP l | T (13) = 3.92 | < 0.00 | 0.03 | 1.81 |
| PaCiG l | aSTG l | T (13) = 3.78 | < 0.00 | 0.03 | 1.75 |
| PaCiG l | aSTG r | T (13) = 5.06 | < 0.00 | 0.03 / 0.03* | 2.34 |
| PaCiG l | HG l | T (13) = 3.67 | < 0.00 | 0.03 | 1.70 |
| PaCiG l | PT l | T (13) = 4.35 | < 0.00 | 0.03 | 2.01 |
| PaCiG l | pMTG l | T (13) = 3.85 | < 0.00 | 0.03 | 1.78 |
| PaCiG l | pSTG l | T (13) = 4.44 | < 0.00 | 0.03 | 2.05 |
| PaCiG l | toMTG l | T (13) = 3.47 | < 0.00 | 0.04 | 1.60 |
| PaCiG l | LG l | T (13) = 3.84 | < 0.00 | 0.03 | 1.78 |
| PaCiG l | LG r | T (13) = 3.28 | 0.01 | 0.05 | 1.51 |
| PaCiG l | ICC l | T (13) = 3.78 | < 0.00 | 0.03 | 1.74 |
| PaCiG l | ICC r | T (13) = 3.65 | < 0.00 | 0.03 | 1.69 |
| PaCiG l | SCC l | T (13) = 3.59 | < 0.00 | 0.03 | 1.66 |
| PaCiG l | SCC r | T (13) = 3.91 | < 0.00 | 0.03 | 1.81 |
| PaCiG l | Cereb45 r | T (13) = 4.10 | < 0.00 | 0.03 | 1.89 |
| PaCiG l | Cuneal r | T (13) = 3.83 | < 0.00 | 0.03 | 1.77 |
| PaCiG r | Cereb45 r | T (13) = 5.24 | < 0.00 | 0.02 | 2.42 |
| aSTG r | PreCG r | T (13) = 4.51 | < 0.00 | 0.04 | 2.08 |
| aTFusC r | aSMG r | T (13) = 5.20 | < 0.00 | 0.02 / 0.02* | 2.40 |
| Cereb45 r | PaCiG r | T (13) = 5.24 | < 0.00 | 0.02 | 2.42 |

*indicates connections which were statistically significant in both the directions (i.e., from Region A – Region B and Region B – Region A): the first $p$-value is for the reported connection and the second $p$-value is for the connection in the reverse direction

#### Within-network connectivity

##### *CAT (n = 15) > HS (n = 15)*

The within network connectivity differences for CAT > HS contrast are shown in **Figure 6** and statistics are reported in **Table 10**. Of these, the sensorimotor, salience and cerebellar networks showed reduced within-network connectivity even after Bonferroni correction for the number of between-group comparisons made ($n$ = 5 networks; $p$-FDR <0.01). The sensorimotor network retained an excellent Dice coefficient even at the Bonferroni-corrected significance threshold (0.92 ± 0.15) indicating 'high' reliability, while the Dice co-efficient for the other networks were not satisfactory (see **Figure 7** for connections within each network which survived Bonferroni correction for multiple comparisons and see **Figure 8** for Dice coefficient for each network on jackknife reliability analysis before and after Bonferroni correction). After jackknife analysis, we found the following connections to be consistently significantly different between groups across all jackknife samples: all connections in the sensorimotor network; connections between left and right anterior insula, left anterior insula and left supramarginal gyrus, and between right and left anterior insula (for salience network); connections between right posterior parietal cortex and left lateral prefrontal cortex, and right and left posterior parietal cortex (for frontoparietal network); and connections between left cerebellum 3 and right cerebellum 9, right cerebellum 9 and left cerebellum 3, and right cerebellum 9 and right cerebellum 3 (for cerebellar network). After Bonferroni correction for the five within-network comparisons between the groups, the following pairs of connections were still consistently significantly different across all jackknife samples: lateral right to lateral left, lateral right to superior, and superior to lateral right (within sensorimotor network); these results are presented in **Figure 7**. See **Figure 8** for Dice coefficient for each network on jackknife reliability analysis before and after Bonferroni correction.

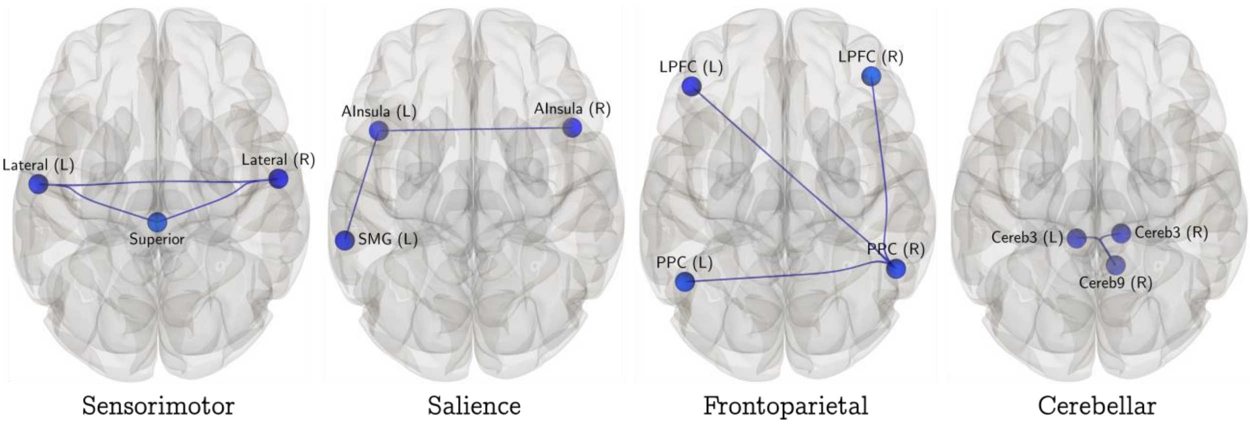

**Figure 6:** Comparison of within-network ROI-ROI connectivity between catatonia group ($n$ = 15) and healthy group ($n$ = 15) (CAT > HS contrast) for sensorimotor, salience, frontoparietal, and cerebellar network at seed-level $p$-FDR < 0.05 threshold for each network; Fisher transformed correlation coefficient between the haemodynamic-response-function-weighted mean regional time series was used as a measure of connectivity; left hemisphere regions are suffixed with '(L)' and right hemisphere regions are suffixed with '(R)'; AInsula: anterior insula; SMG: supramarginal gyrus; LPFC: lateral prefrontal cortex; PPC: posterior parietal cortex

**Table 10:** Pairs of connections within the sensorimotor, salience, frontoparietal, and cerebellar networks that were significantly different between catatonia group ($n$ = 15) and healthy group ($n$ = 15) (CAT > HS contrast) at seed-level $p$-FDR < 0.05 threshold for each network; all connections were in the negative direction i.e. reduced in the catatonia group as compared to the healthy group; connections which were statistically significant in both directions have two values in the $p$-FDR column; $p$-values are rounded off to two decimal places; the $p$-FDR values of connections which survived an additional Bonferroni correction for five networks are underlined; left and right sides are indicated with 'l' and 'r' suffix

| Source | Target | *T* (df) Statistics | *p*-uncorrected | *p*-FDR | Hedges' *g* |
|---|---|---|---|---|---|
| Sensorimotor network | | | | | |
| Lateral l | Lateral r | T (28) = -3.52 | < 0.00 | < 0.00 / < 0.00* | 1.21 |
| Lateral l | Superior | T (28) = -3.14 | < 0.00 | < 0.00 / < 0.00* | 1.08 |
| Lateral r | Superior | T (28) = -3.99 | < 0.00 | < 0.00 / < 0.00* | 1.37 |
| Salience network | | | | | |
| AInsula l | SMG l | T (28) = -3.22 | < 0.00 | 0.01 / 0.02* | 1.10 |
| Ainsula l | Ainsula r | T (28) = -3.57 | < 0.00 | 0.01 / 0.01* | 1.22 |
| Frontoparietal network | | | | | |
| LPFC l | PPC r | T (28) = -2.93 | 0.01 | 0.02 / 0.01* | 1.01 |
| PPC l | PPC r | T (28) = -2.90 | 0.01 | 0.02 / 0.01* | 1.00 |
| PPC r | LPFC r | T (28) = -2.40 | 0.02 | 0.02 | 0.82 |
| Cerebellar network | | | | | |
| Cereb3 l | Cereb9 r | T (28) = -4.23 | < 0.00 | 0.01 / 0.01* | 1.45 |
| Cereb3 r | Cereb9 r | T (28) = -3.78 | < 0.00 | 0.02 / 0.01* | 1.30 |

AInsula: anterior insula; SMG: supramarginal gyrus; LPFC: lateral prefrontal cortex; PPC: posterior parietal cortex; *indicates connections which were statistically significant in both the directions (i.e., from Region A – Region B and Region B – Region A): the first *p*-value is for the reported connection and the second *p*-value is for the connection in the reverse direction

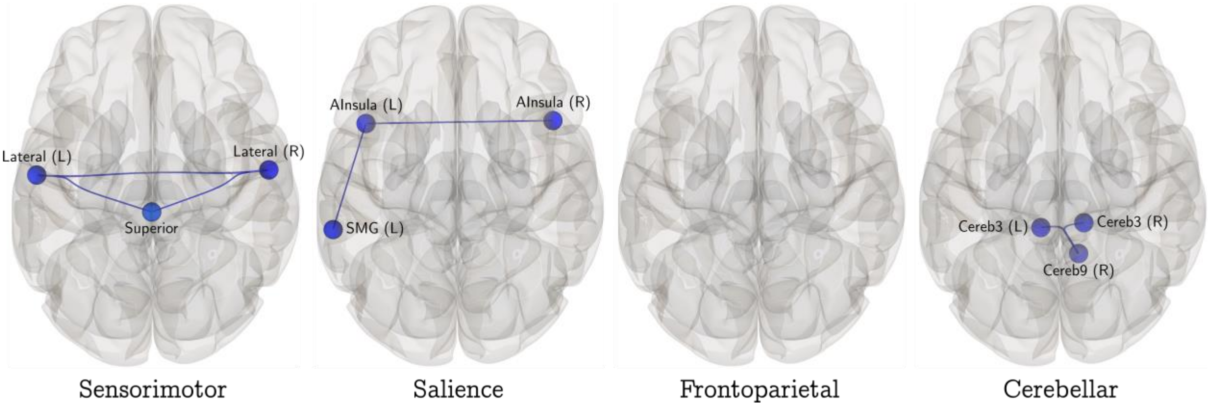

**Figure 7:** Comparison of within-network ROI-ROI connectivity between catatonia group (*n* = 15) and healthy group (*n* = 15) (CAT > HS contrast) for sensorimotor, salience, frontoparietal, and cerebellar networks with an additional Bonferroni correction for multiple comparisons (correction for five networks) i.e. at seed-level *p*-FDR < 0.01 threshold for each network; Fisher transformed correlation coefficient between the haemodynamic-response-function-weighted mean regional time series was used as a measure of connectivity; left hemisphere regions are suffixed with '(L)' and right hemisphere regions are suffixed with '(R)'; AInsula: anterior insula; SMG: supramarginal gyrus; LPFC: lateral prefrontal cortex; PPC: posterior parietal cortex

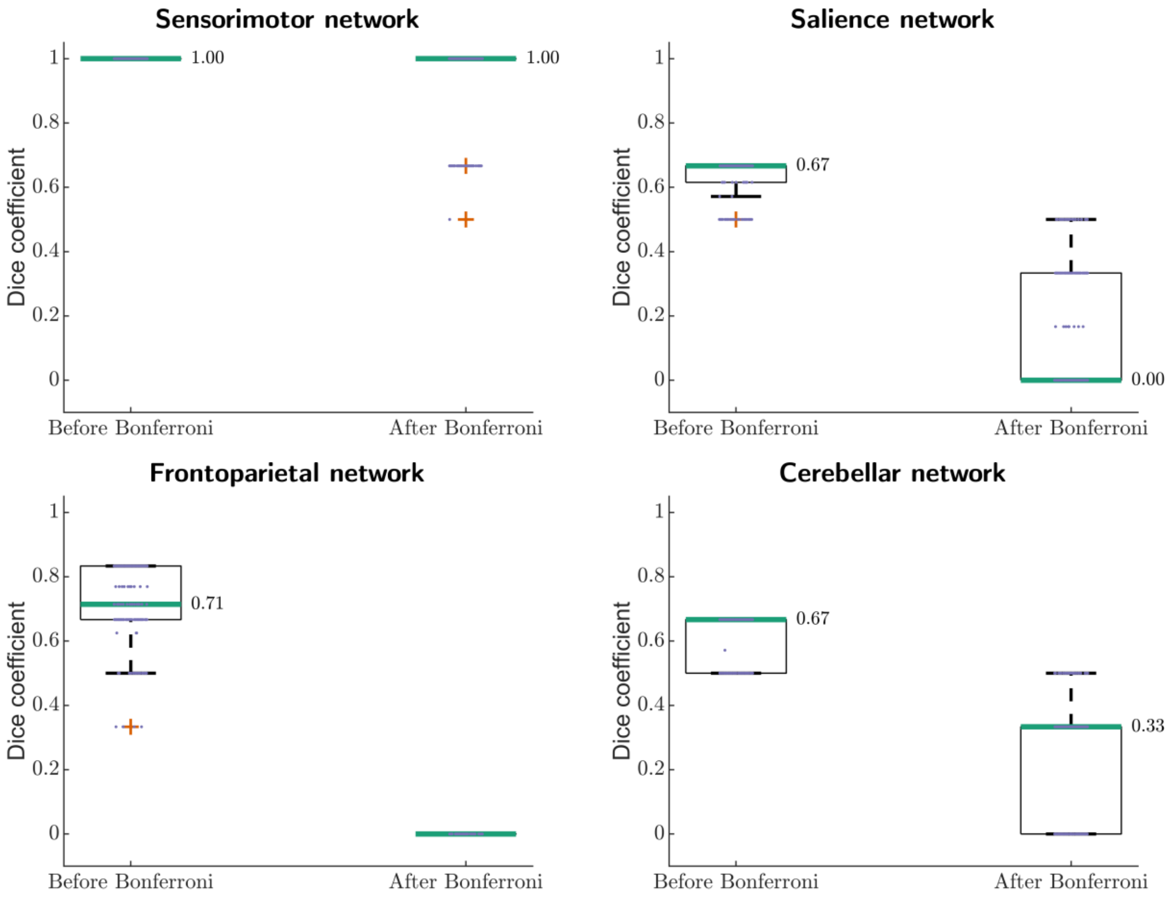

**Figure 8:** Boxplots of Dice coefficients from jackknife analyses for between group comparisons of within network connectivity for sensorimotor, salience, frontoparietal, and cerebellar networks (with and without Bonferroni correction for multiple comparisons) (CAT > HS contrast)

##### *CAT (n = 15) > HS (Achieva only; n = 11)*

When comparing the within network ROI-ROI connectivity for sensorimotor, salience, frontoparietal, cerebellar, and subcortical (basal ganglia) networks (individually) between catatonia ($n$ = 15) and healthy samples (excluding 4 healthy subjects whose images were acquired on Ingenia CX scanner; $n$ = 11) [CAT > HS (Achieva only) contrast], we found reduced connectivity in the catatonia sample within sensorimotor, salience, frontoparietal, and cerebellar networks. We did not find any statistically significant differences within the subcortical network between the two groups. These results were similar to the case of considering all healthy subjects together. Results for the sensorimotor, salience, frontoparietal, and cerebellar networks are shown in **Figure 9** and statistics are reported in **Table 11**.

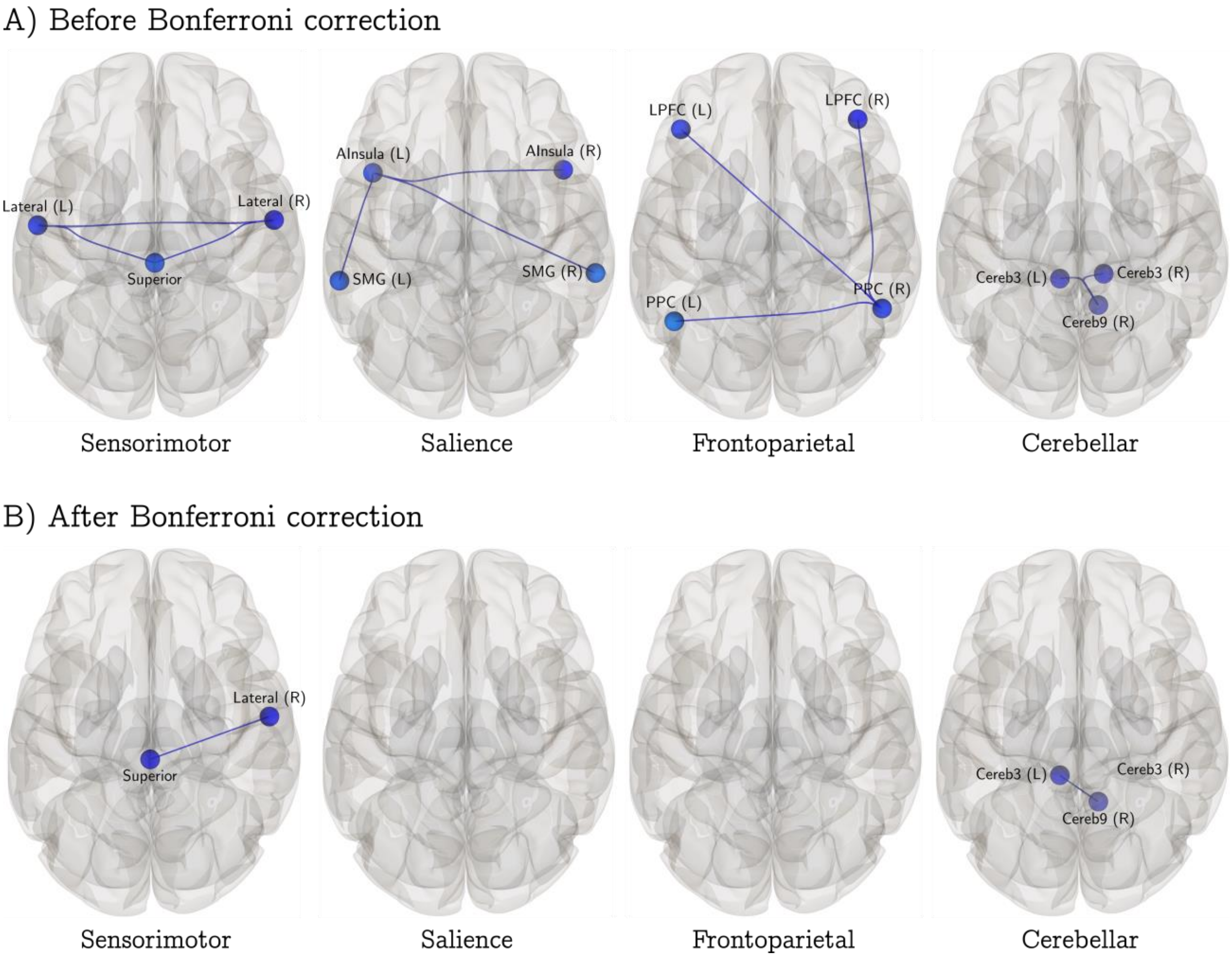

**Figure 9:** Comparison of within network ROI-ROI connectivity between catatonia group ($n$ = 15) and healthy group (excluding those images which were acquired on Ingenia CX scanner; $n$ = 11), CAT > HS (Achieva only) contrast for sensorimotor, salience, frontoparietal, and cerebellar network at **A)** seed-level $p$-FDR < 0.05 threshold for each network; and **B)** with an additional Bonferroni correction for five networks; Fisher transformed correlation coefficient between the haemodynamic response function weighted mean regional time series was used as a measure of connectivity; left hemisphere regions are suffixed with '(L)' and right hemisphere regions are suffixed with '(R)'; AInsula: anterior insula; SMG: supramarginal gyrus; LPFC: lateral prefrontal cortex; PPC: posterior parietal cortex

**Table 11:** Pairs of connections within the sensorimotor, salience, frontoparietal, and cerebellar networks that were significantly different between catatonia group ($n$ = 15) and healthy group (excluding those images which were acquired on Ingenia CX scanner; $n$ = 11), CAT > HS (Achieva only) contrast at seed-level $p$-FDR < 0.05 threshold for each network; all connections were in the negative direction i.e. reduced in the catatonia group as compared to the healthy group; the connections are ordered from anterior to posterior source ROIs, followed by subcortical source ROIs, and finally by cerebellar source ROIs; connections which were statistically significant in both directions have two values in the $p$-FDR column; $p$-values are rounded off to two decimal places; AInsula: anterior insula; SMG: supramarginal gyrus; LPFC: lateral prefrontal cortex; PPC: posterior parietal cortex; the $p$-FDR values of connections which survived an additional Bonferroni correction for five networks are underlined

| Source | Target | *T* (df) Statistics | *p*-uncorrected | *p*-FDR |
|---|---|---|---|---|
| Sensorimotor network | | | | |
| Left Lateral | Right Lateral | T (24) = -2.60 | 0.02 | 0.03 / 0.02* |
| Left Lateral | Superior | T (24) = -2.34 | 0.03 | 0.03 / 0.03* |

| | | | | |
|---|---|---|---|---|
| Right Lateral | Superior | T (24) = -3.11 | < 0.00 | 0.01 / 0.01* |
| Salience network | | | | |
| Left AInsula | Left SMG | T (24) = -2.65 | 0.01 | 0.04 |
| Left AInsula | Right AInsula | T (24) = -3.53 | < 0.00 | 0.01 / 0.01* |
| Right AInsula | Left AInsula | T (24) = -3.53 | < 0.00 | 0.01 |
| Frontoparietal network | | | | |
| Right LPFC | Right PPC | T (24) = -2.61 | 0.02 | 0.05 / 0.04* |
| Right PPC | Left LPFC | T (24) = -2.41 | 0.02 | 0.04 |
| Right PPC | Left PPC | T (24) = -2.16 | 0.04 | 0.04 |
| Cerebellar network | | | | |
| Cereb3 l | Cereb9 r | T (24) = -4.32 | < 0.00 | 0.01 / 0.01* |
| Cereb3 r | Cereb9 r | T (24) = -3.78 | < 0.00 | 0.02 / 0.01* |

*indicates connections which were statistically significant in both the directions (i.e. from Region A – Region B and Region B – Region A): the first *p*-value is for the reported connection and the second *p*-value is for the connection in the reverse direction

#### Aberrant functional connectivity of the motor cortex in acute catatonia

##### *CAT (n = 15) > HS (n = 15)*

The results of seed (left precentral gyrus)-to voxel connectivity analysis are presented in **Figure 10 a**nd the brain regions covered by the significant clusters are summarized in **Figure 10b**; cluster size and cluster-wise *p*-values are reported in **Table 12**.

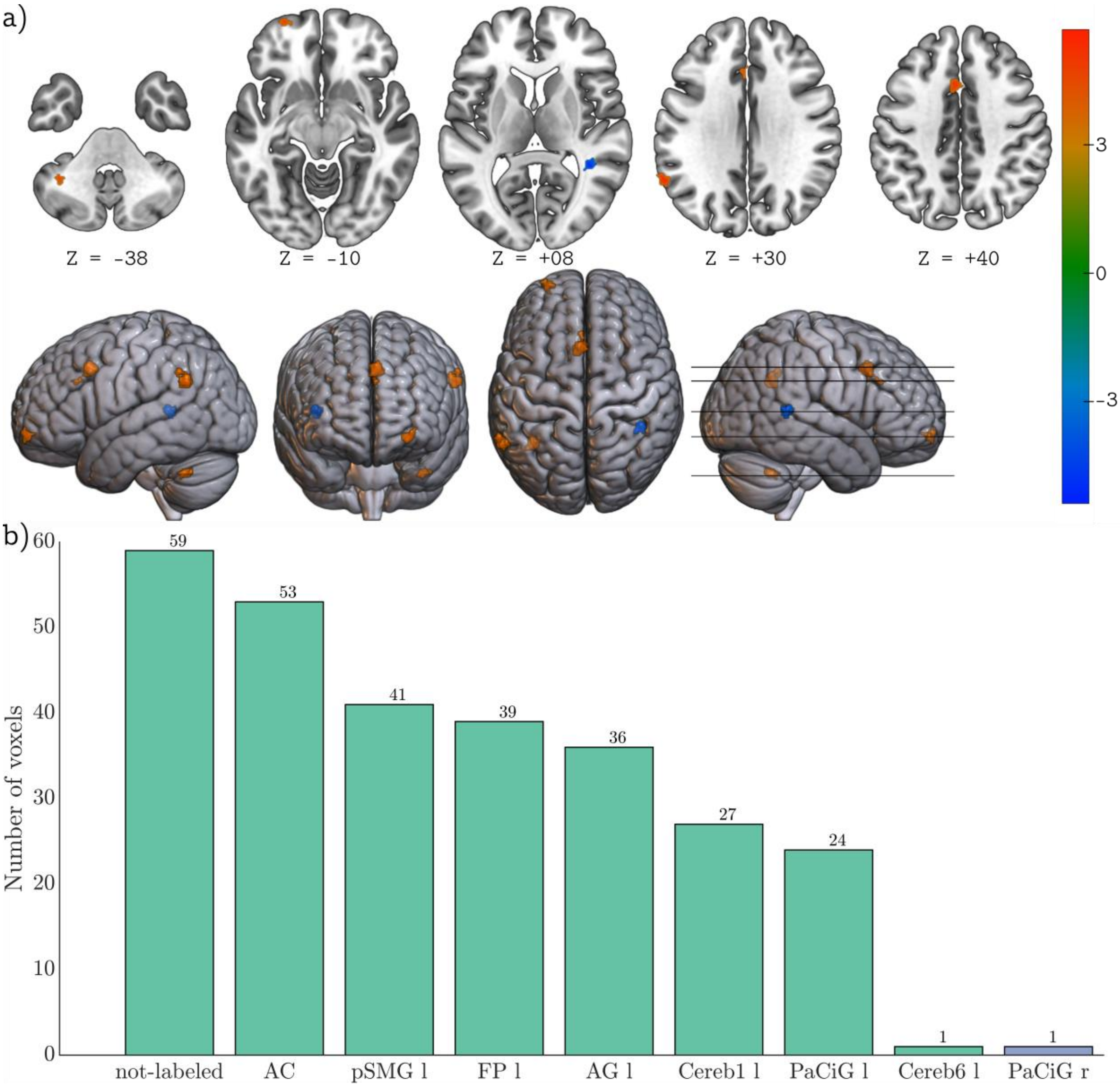

**Figure 10:** Comparison of seed-to-voxel connectivity from the left precentral gyrus between catatonia group ($n$ = 15) and the healthy group ($n$ = 15) (CAT > HS contrast); **a)** clusters with significantly different mean connectivity between the groups with blue colors indicating reduced connectivity in the catatonia group (unlabeled voxels) and red colors indicating increased connectivity in the catatonia group, compared to healthy group at voxel-wise uncorrected $p < 0.001$, cluster-wise FDR corrected $p < 0.05$ ($T$min = 3.67, $k$min = 39); color bar range is from -5.378 to 5.683; **b)** list of regions and the number of voxels within these regions covered by these clusters; the left pane shows regions from the left hemisphere, the regions which are not split by hemisphere, and the unlabeled voxels, while the right pane shows regions from the right hemisphere; see **Table 5** for expansion of the abbreviations used for the brain regions

**Table 12:** Clusters showing significantly different left precentral gyrus-based connectivity between catatonia group ($n$ = 15) and healthy group ($n$ = 15), CAT > HS contrast, at voxel-wise uncorrected $p < 0.001$, cluster-wise FDR corrected $p < 0.05$ ($T$min = 3.67, $k$min = 39); all $p$-values are rounded to two decimal places; FWE: family-wise error; FDR: false discovery rate

| Cluster (x, y, z) | Size | Size $p$-FWE | Size $p$-FDR | Size $p$-uncorrected | Peak $p$-FWE | Peak $p$-uncorrected | Hedges' $g$ |
|---|---|---|---|---|---|---|---|
| -04 +16 +40 | 82 | < 0.00 | < 0.00 | < 0.00 | 0.72 | < 0.00 | 1.92 |
| -58 -54 +30 | 78 | 0.01 | < 0.00 | < 0.00 | 0.63 | < 0.00 | 1.96 |

| | | | | | | | |
|---|---|---|---|---|---|---|---|
| +40 -44 +08 | 42 | 0.13 | 0.05 | < 0.00 | 0.72 | < 0.00 | 1.93 |
| -36 -52 -38 | 40 | 0.16 | 0.05 | < 0.00 | 0.99 | < 0.00 | 1.72 |
| -26 +60 -10 | 39 | 0.17 | 0.05 | < 0.00 | 0.92 | < 0.00 | 1.82 |

##### *CAT (n = 15) > HS (Achieva only; n = 11)*

When comparing the left precentral gyrus seed-based connectivity between the catatonia group ($n$ = 15) and the healthy group (excluding 4 healthy subjects whose images were acquired on Ingenia CX scanner; $n$ = 11) [CAT > HS (Achieva only) contrast], we found three clusters of significantly increased connectivity in the catatonia group. These results are presented in **Figure 11a,** and the list of brain regions covered by these clusters is summarized in **Figure 11b**; cluster size and cluster-wise $p$-values are reported in **Table 13**.

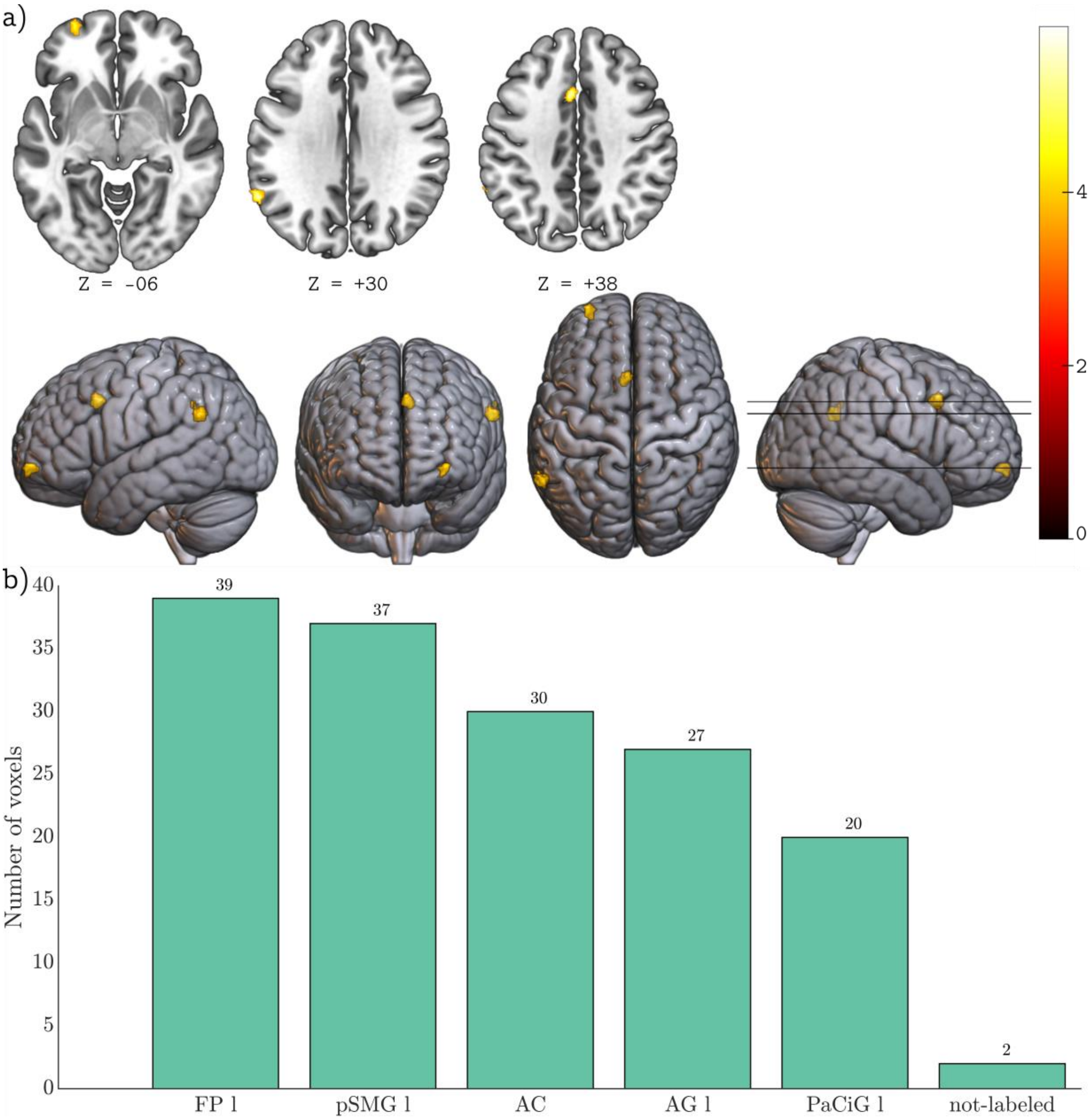

**Figure 11:** Comparison of seed-to-voxel connectivity from the left precentral gyrus between catatonia group ($n$ = 15) and the healthy group (excluding those images which were acquired on Ingenia CX scanner; $n$ = 11), CAT > HS (Achieva only) contrast; **a)** clusters with significantly increased mean connectivity in the catatonia group

compared to healthy group at voxel-wise uncorrected $p < 0.001$, cluster-wise FDR corrected $p < 0.05$ (*T*min = 3.75, *k*min = 39); color bar range is from 0 to 5.908; **b)** list of regions and the number of voxels within these regions covered by these clusters; see **Table 5** for expansion of the abbreviations used for the brain regions

**Table 13:** Clusters showing significantly different seed-to-voxel connectivity from the left precentral gyrus between catatonia group ($n = 15$) and healthy group (excluding those images which were acquired on Ingenia CX scanner; $n = 11$), CAT > HS (Achieva only) contrast at voxel-wise uncorrected $p < 0.001$, cluster-wise FDR corrected $p < 0.05$ (*T*min = 3.75, *k*min = 39); all *p*-values are rounded to two decimal places; FWE: family-wise error; FDR: false discovery rate

| Cluster (x, y, z) | Size | Size *p*-FWE | Size *p*-FDR | Size *p*-uncorrected | Peak *p*-FWE | Peak *p*-uncorrected |
|---|---|---|---|---|---|---|
| -58 -54 +30 | 66 | 0.01 | 0.01 | < 0.00 | 0.70 | < 0.00 |
| -04 +16 +38 | 50 | 0.04 | 0.01 | < 0.00 | 0.86 | < 0.00 |
| -28 +60 -06 | 39 | 0.12 | 0.03 | < 0.00 | 1.00 | < 0.00 |

##### *LZM (n = 9) > ECT (n = 6)*

The left precentral gyrus seed-to-voxel connectivity differences between the lorazepam responder group ($n = 9$) and non-responder group ($n = 6$) (LZM > ECT contrast) are presented in **Figure 12a** and the list of brain regions covered by these clusters is summarized in **Figure 12b**; cluster size and cluster-wise *p*-values are reported in **Table 14**.

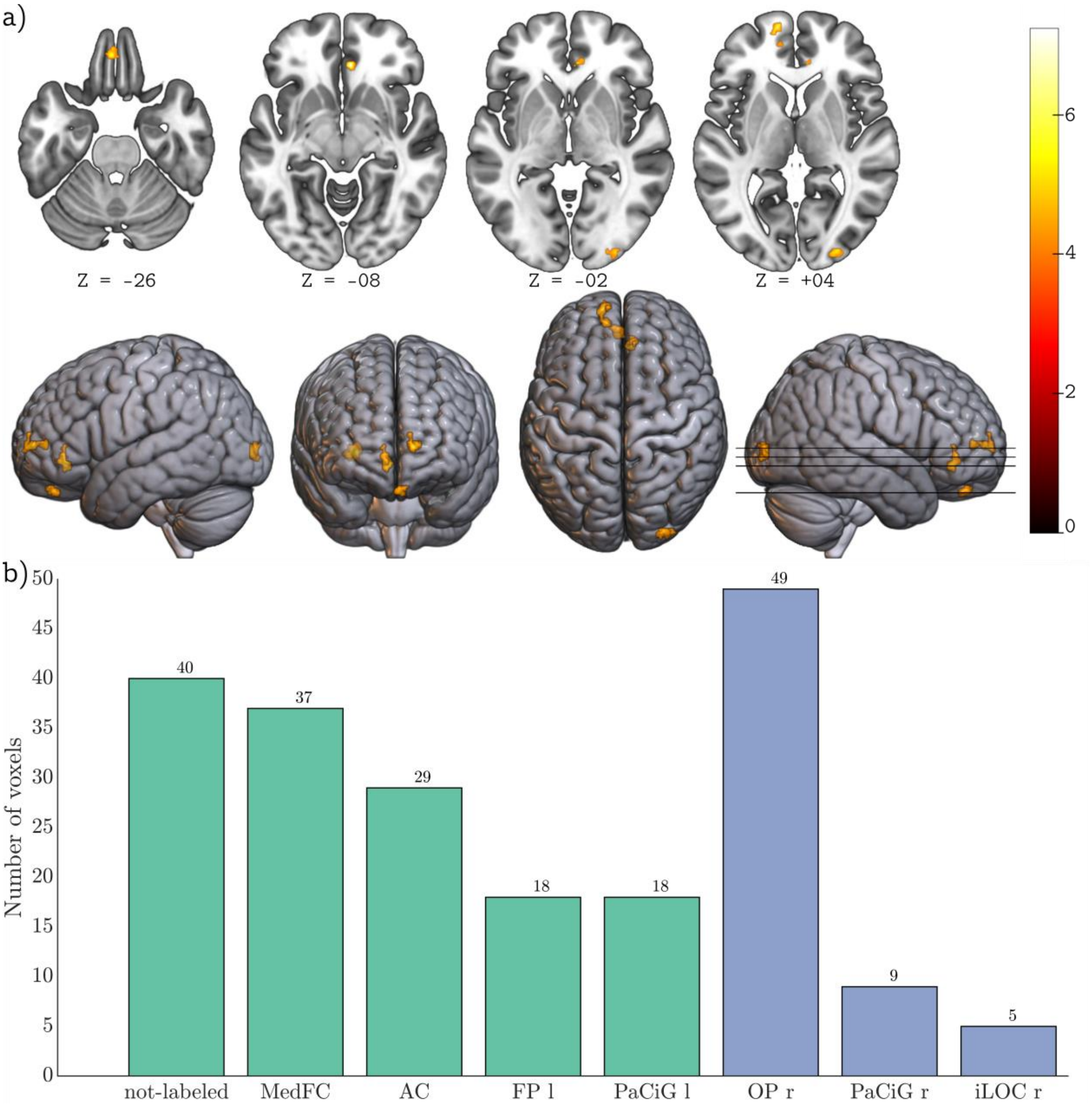

**Figure 12:** Comparison of the seed-to-voxel connectivity from the left precentral gyrus between lorazepam responder group ($n$ = 9) and the lorazepam non-responder group ($n$ = 6) (LZM > ECT contrast); **a)** clusters with significantly increased mean connectivity in the lorazepam responder group compared to lorazepam non-responder group at voxel-wise uncorrected $p < 0.001$, cluster-wise FDR corrected $p < 0.05$ ($T$min = 4.22, $k$min = 35); color bar range is from 0 to 7.238; **b)** list of regions and the number of voxels within these regions covered by these clusters; the left pane shows regions from the left hemisphere, the regions which are not split by hemisphere, and the unlabeled voxels, while the right pane shows regions from the right hemisphere; see **Table 5** for expansion of the abbreviations used for the brain regions

**Table 14:** Clusters showing significantly different seed-to-voxel connectivity from the left precentral gyrus between lorazepam responder group ($n$ = 9) and lorazepam non-responder group ($n$ = 6), LZM > ECT contrast, at voxel-wise uncorrected $p < 0.001$, cluster-wise FDR corrected $p < 0.05$ ($T$min = 4.22, $k$min = 35); all $p$-values are rounded to two decimal places; FWE: family-wise error; FDR: false discovery rate

| Cluster (x, y, z) | Size | Size *p*-FWE | Size *p*-FDR | Size *p*-uncorrected | Peak *p*-FWE | Peak *p*-uncorrected | Hedges' *g* |
|---|---|---|---|---|---|---|---|
| -12 +60 +04 | 60 | < 0.00 | < 0.00 | < 0.00 | 0.94 | < 0.00 | 3.47 |

| | | | | | | | |
|---|---|---|---|---|---|---|---|
| +32 -92 -02 | 60 | < 0.00 | < 0.00 | < 0.00 | 1.00 | < 0.00 | 2.84 |
| +06 +34 -08 | 50 | 0.01 | < 0.00 | < 0.00 | 0.88 | < 0.00 | 3.53 |
| -02 +42 -26 | 35 | 0.05 | 0.01 | < 0.00 | 1.00 | < 0.00 | 2.69 |

##### *Relationship between BFCRS motor sub-score and seed-to-voxel connectivity from left precentral gyrus*

In this regression analysis, the BFCRS motor sub-score was used as a predictor of seed-to-voxel connectivity from the left precentral gyrus; these results are presented in **Figure 13a** and the brain regions covered by the cluster are presented in **Figure 13b**.

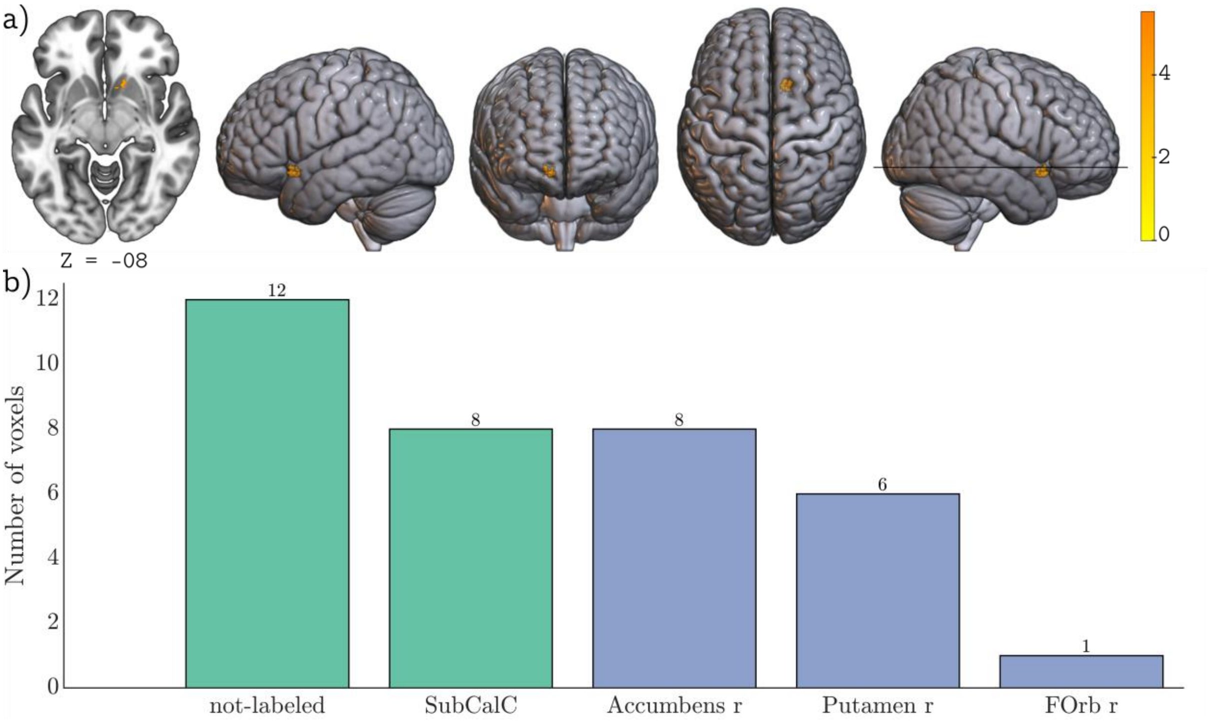

**Figure 13:** Regression analysis between BFCRS motor sub-scores and seed-to-voxel connectivity from the left precentral gyrus in the catatonia group ($n$ = 15); **a)** clusters where connectivity was significantly predicted at voxel-wise uncorrected $p < 0.001$, cluster-wise FDR corrected $p < 0.05$ ($T$min = 4.22, $k$min = 35); color bar range is from 0 to 5.524; **b)** list of regions and the number of voxels within these regions covered by this cluster; the left pane shows regions from the left hemisphere, the regions which are not split by hemisphere, and the unlabeled voxels, while the right pane shows regions from the right hemisphere; see **Table 5** for expansion of the abbreviations used for the brain regions

#### Altered cortical complexity in catatonia

##### *HS (n = 15) > CAT (n = 15)*

The cortical complexity differences between the healthy and catatonia groups are shown in **Figure 14**.

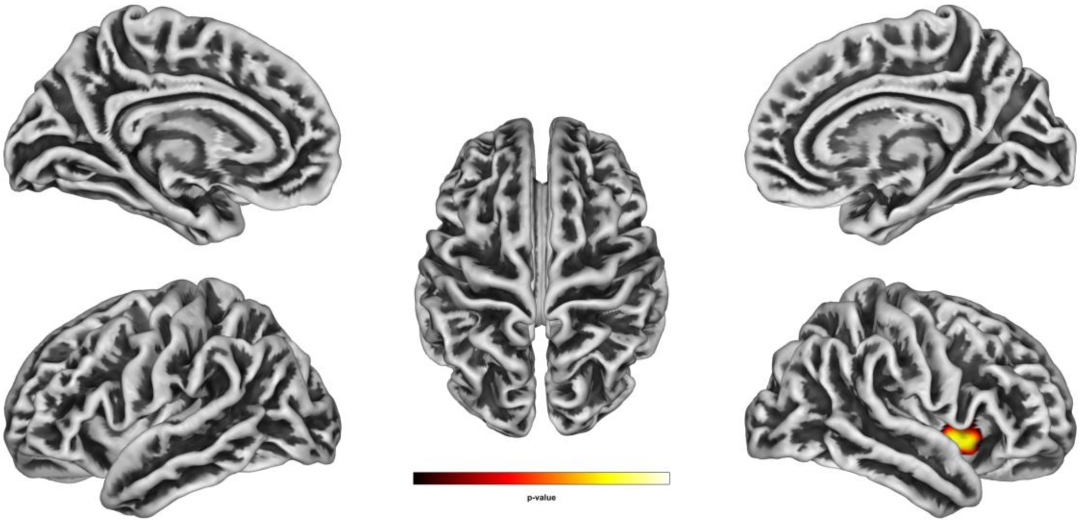

**Figure 14:** Comparison of vertex-wise cortical complexity between healthy group (*n* = 15) and catatonia group (*n* = 15) at a threshold of $p < 0.05$ (FWE corrected) using a non-parametric TFCE approach; ; HS > CAT contrast

##### *HS (Achieva only; n = 11) > CAT (n = 15)*

On comparing the cortical complexity between healthy subjects (excluding 4 healthy subjects whose images were acquired on Ingenia CX scanner) and catatonia patients, we found two clusters of reduced cortical complexity in the patient group. These results are presented in **Figure 15** and the statistics and lookup information are presented in **Table 15**.

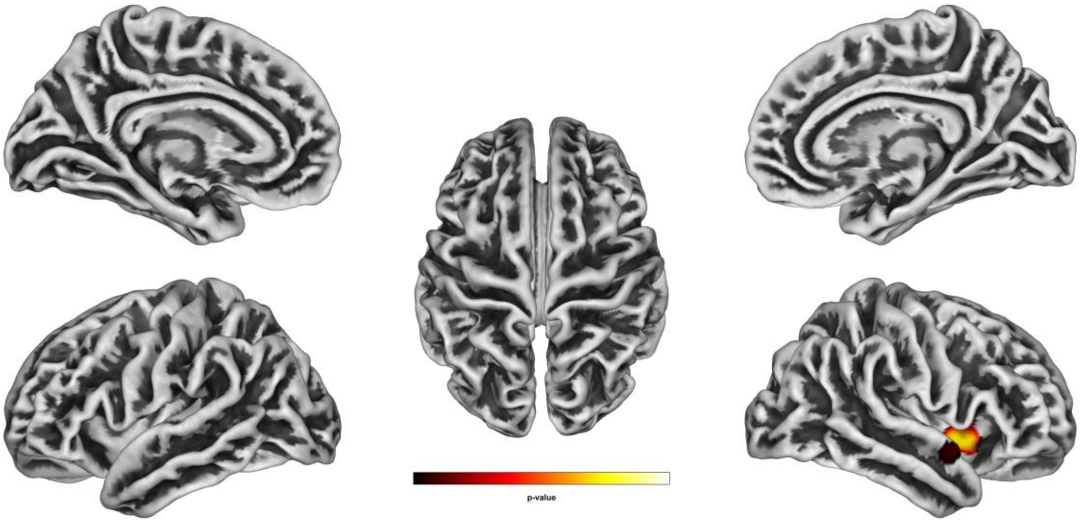

**Figure 15:** Comparison of vertex-wise cortical complexity between healthy group (excluding those images which were acquired on Ingenia CX scanner; *n* = 11) and catatonia group (*n* = 15) at a threshold of $p < 0.05$ (FWE corrected) using a non-parametric TFCE approach; HS > CAT contrast

**Table 15:** Clusters of significantly different cortical complexity between healthy group (excluding the images which were acquired on Ingena CX scanner; $n = 11$) and catatonia group ($n = 15$) at a threshold of $p < 0.05$ (FWE corrected) using a non-parametric TFCE approach; HS > CAT contrast

| *p*-value | Size | Overlap | Region (Right hemisphere) |
|---|---|---|---|
| 0.02 | 274 | 34% | Middle insular area |
| | | 20% | Posterior insular area 2 |
| | | 18% | Anterior agranular insular area |
| | | 13% | Frontal opercular area 3 |
| | | 7% | Piriform cortex |
| | | 7% | Posterior insular area 1 |
| | | 1% | Frontal opercular area 4 |
| 0.04 | 45 | 56% | Dorsal superior temporal sulcus |
| | | 42% | Auditory complex 5 |
| | | 2% | Temporal region A2 |

### Strengths and limitations of the study

The novel findings that emerge from this first fMRI study in acute retarded catatonia carried out in an emergency psychiatry setting, substantially advances our understanding of the underlying neurobiology of catatonia and its response to benzodiazepines. Given the challenges involved in carrying out such a study, the sample sizes of the catatonia group and especially of the lorazepam responder and non-responder subgroups were modest. While this may be considered a limitation, it is noteworthy that a clear-cut differentiation with effect sizes ranging from high (0.8) to huge (>2) Hedges' *g* values[30,31] between catatonia and healthy samples as well as between lorazepam responder and non-responder groups was observed at a fairly stringent statistical threshold in this modest sample, indicating the robust nature of these functional brain abnormalities. Furthermore, we have carried out additional reliability analyses using a jackknife approach which showed 'perfect' (within-network connectivity reduction in the sensorimotor network), 'high' (within-network connectivity reduction in the frontoparietal network) and fair (increased whole brain rsFC; within-network connectivity reduction in the salience and cerebellar networks) Dice coefficients, indicating good reliability of our main results (the qualifiers 'perfect' and 'high' are based on[28]). This initial fMRI study in acute retarded catatonia was carried out in patients who were in the acute catatonic state at the time of MRI acquisition as ascertained by baseline administration of the Bush Francis Rating Scale within 1 hour prior to the start of the scanning session. We made our best efforts to acquire the MRI scans prior to initiation of treatment with lorazepam; however, in eight out of the 15 patients with catatonia, we could scan the patient only after initiation of treatment with lorazepam (see Table 3) due to practical issues related to scanner availability at short notice. Nevertheless, the mean BFCRS rating at baseline which was performed within an hour prior to the MRI was 21.07 (standard deviation = 5.69; minimum = 12; maximum = 31), indicating that all participants were in acute catatonia at the time of scanning.
